# Forecasting the Spread of the COVID-19 Epidemic in Lombardy: A Dynamic Model Averaging Approach

**DOI:** 10.1101/2021.01.18.21250053

**Authors:** Lisa Gianmoena, Vicente Rios

**Affiliations:** Department of Economics and Management, University of Pisa, Cosimo Ridolfi 10, 56124, Pisa, Italy.; Department of Economics, Management and Quantitative Methods, University of Milan, Via Festa del Perdono 7 - 20122 Milano, Italy

**Keywords:** COVID-19 epidemic, Lombardy, Real-time Forecasts, Dynamic Model Averaging

## Abstract

Forecasting with accuracy the evolution of COVID-19 daily incidence curves is one of the most important exercises in the field of epidemic modeling. We examine the forecastability of daily COVID-19 cases in the Italian region of Lombardy using Dynamic Model Averaging and Dynamic Model Selection methods. To investigate the predictive accuracy of this approach, we compute forecast performance metrics of sequential out-of-sample real-time forecasts in a back-testing exercise ranging from March 1 to December 10 of 2020. We find that (i) Dynamic Model Averaging leads to a consistent and substantial predictive improvements over alternative epidemiological models and machine learning approaches when producing short-run forecasts. Using estimated posterior inclusion probabilities we also provide evidence on which set of predictors are relevant for forecasting in each period. Our findings also suggest that (ii) future incidences can be forecasted by exploiting information on the epidemic dynamics of neighboring regions, human mobility patterns, pollution and temperatures levels.

## 1 Introduction

The Coronavirus Disease 2019 (COVID-19) pandemic produced by the Severe Acute Respiratory Syndrome Corona Virus (SARS-CoV-2) pathogen is likely to have been the most disruptive shock to our societal organization since the World War II, threatening both, the health systems and the functioning of the economy (WHO, 2020; IMF, 2020). In this regard, during the health-crisis posed by the COVID-19, one of the most relevant and pervasive problems from a policy-making point of view has been the inability to anticipate with accuracy the evolution of the epidemic and its pressure exerted on health-systems (Ioannidis *et al*., 2020). The negative consequence of these failed forecasts (either over-pessimistic or over optimistic) has been a reduced government’s ability to implement the required policies in time.

By the end of 2020, most of the world population lacks of immunity and remains susceptible to the disease, making public health officials to be concerned with the threat of future COVID-19 outbreaks and about the severity of future waves. In this context, forecasting with accuracy the evolution of incidence curves is one of the most important exercises in the field of epidemic modeling and forecasting. This is because of long-term regional epidemic forecasts (i.e, months to years) of the COVID-19 pandemic can be useful to make strategic decisions regarding the location and number of testing, treatment facilities, or the distribution of the vaccines. On the other hand, short-term forecasts (i.e, days to weeks) can be helpful to anticipate resources such as protective equipment, medical ventilators, hospital beds or take the decisions an aid on the implementation timing of lock-downs and restrictions (Chowell *et al*., 2020).

Epidemiologists have commonly used *compartmental models* to forecast the expected epidemic disease trajectories being the most widely used one the Susceptible-Infected-Recovered (SIR) (Kermack and McKrendrick, 1927). In the context of the COVID-19, early SIR applications (and its variants such as the Susceptible-Exposed-Infected-Recovered (SEIR) and Susceptible-Infected-Recovered-Death (SIRD)) to forecast the evolution of contagion and deaths can be found in Roda *et al*. (2020), Anastassopoulou *et al*. (2020) and Fanelli and Piazza (2020) among others. These models are based on systems of ordinary differential equations and focus on the dynamic progression of a population through different epidemiological states. However, an important drawback of compartmental models is that as their complexity increases (i.e, new states are modeled), the stronger the problem of parameter identification becomes, which can deteriorate their forecasting performance (Korolev, 2020). In fact, as shown by (Roda *et al*., 2020), in the context of the COVID-19 outbreak, predictions from complex models might not be so reliable when compared to those of simpler ones. For this reason, other strand of mathematical epidemic modelers have employed to more parsimonious and simpler *phenomenological models* of epidemic growth (Roosa *et al*., 2020a,b) to forecast the evolution of incidence curves.^1^

In either case, there are critical issues that these workhorse epidemiological models fail to account for.

First, these modeling frameworks are silent on the role played by exogenous factors and usually neglect the effect of model uncertainty in their predictions. However, the implementation of lock-downs (Born *et al*., 2020; Deb *et al*., 2020), the changing climate (**?**; Paez *et al*., 2020; Rios and Gianmoena, 2020) and pollution patterns (Sciomer *et al*., 2020; Yongjian *et al*., 2020; Wu *et al*., 2020), the restrictions on social mobility within and across regions (Cartení et al., 2020; Kraemer *et al*., 2020; Zhou *et al*., 2020) or the laws on the use of protective equipment such as face masks or distancing measures (Mitze *et al*., 2020; Wang, Y. *et al*., 2020a), are likely to have affected the spread dynamics of the COVID-19. Given that it is not clear which set of factors could be part of the data generating process, a naive approach that ignores model uncertainty may result in biased estimates, overconfident (too narrow) standard errors and misleading inference and predictions.^2^ In fact, when considering a set of *K* potential predictors of incidence, researchers face a large model space formed by *k* = 1, …, ^*K*^ forecasting models *M*_*k*_. This contrasts with the common practice in the field of epidemic modeling of exploiting information on the links between few population variables and their past values within a single model framework.

Second, the forecasting models of incidences might be subject to structural breaks and other sources of parameter instability. Hence, the influence that different variables or predictors could exert on contagions might be time-varying. This feature of epidemic processes may be addressed by means of time-varying parameter models (TVPs), but these techniques are not commonly employed in epidemic analysis.^3^ Kraemer *et al*. (2020) shows the relevance of this point by using real-time mobility data from Wuhan finding that mobility played a large role in the spread of the virus initially but after the implementation of control measures, the correlation between infection growth rates and mobility dropped significantly.

Finally, a problematic issue that needs to be accounted for when forecasting COVID-19 is that the best forecasting model at time 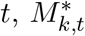, can quickly become obsolete due to rapid changes in the factors driving transmission rates (i.e, environmental factors, behavioral changes in the population and/or by government interventions). For example, it is possible that the best predictors and models to explain accelerations are different to those that perform well during phases of slowdown. Likewise, it may also be optimal to use many predictors at some points in time but only a few of them at others.

Thus, to account for both, (i) the uncertainty regarding the inclusion of the many potential drivers of infections forming model specifications at each date and (ii) the variation over time of the parameters when forecasting the spread of COVID-19, we employ Dynamic Model Averaging (DMA) and Dynamic Model Selection (DMS) methods developed by Raftery *et al*. (2010) and popularized by Koop and Korobilis (2012) in the field of macro-econometrics. DMA/DMS approaches have some advantages with respect current epidemic modeling and forecasting frameworks. In DMA, the weight of a model in a particular period is directly connected with the model predictive likelihood based on past information, while DMS selects the model with the highest probability at each time. Thus, the DMA or DMS approaches seem ideally suited for the problem of COVID-19 forecasting, since they allow for the forecasting model and the coefficients to evolve over time, thereby capturing rapid changes in the effects of the potential determinants COVID-19. Moreover, this data-driven approach involve only standard econometric methods for state space models such as the Kalman filter, while achieving important gains in computational efficiency.

We contribute to the growing literature of epidemic modeling and forecasting by adopting the DMA and DMS frameworks to forecast COVID-19 outcomes. To the best of our knowledge, this is the first study applying DMA and DMS approaches to the context of regional COVID-19 forecasts, and the only one covering the full history of the COVID-19 pandemic and not specific sub-samples. Specifically, we perform an exercise of sequential out-of-sample real-time short-term forecasts using daily incidence data from March 1 to December 10 of 2020. We take the Italian region of Lombardy as our testing ground for two reasons. Lombardy was not only the epicenter of the COVID-19 pandemic in the western world during the first wave, but it has also been one of the European regions that has been hard hit the most by the COVID-19 pandemic during the second wave. This time pattern has been caused by explosive paths and abrupt changes in the transmission of the disease. Hence, the historical epidemic path in Lombardy makes the task of forecasting with accuracy the figures of this region specially challenging. A second reason is that the Italian Civil Protection Ministry provides longer time-series and more reliable and homogeneous data in the key regional magnitudes of the epidemic than other countries (Morettini *et al*., 2020).^4^

Using epidemic data for Lombardy, we show that DMA/DMS methods combining time-varying parameters with the information contained in a large set of models have the potential to improve the forecast accuracy of the new cases series when compared to other competing models used in the fields of epidemiology and machine learning.

## 2 Data

### 2.1 The evolution of COVID-19 incidences in Lombardy

The indicator to capture the dynamics of COVID-19 under scrutiny in this analysis is the time series of new cases or the **daily incidence**, which was collected between February 24, 2020 to December 10, 2020 from the Italian Ministry of Civil Protection (MCP) website. Hence, the considered time series consisted of 291 observations.

Panel (a) of Figure (1), plots the daily incidence curve of Lombardy from Feb 24 to December 10, whereas panel (b) plots the relative share of new cases and cumulative incidences with respect the country’s aggregate. Panel (a) reveals the spread of the COVID-19 in Lombardy has two distinct phases. The first wave covers the period ranging from February 24 to the end of July of 2020. This period is characterized by an explosive growth path until the 21 of March, where it reached a peak with 3,252 cases. After peaking, new incidences experienced a strong and sustained reduction. The second wave spans from August 2020 to December 2020, peaking on November 7 with 11,490 cases. An issue is that the raw data curve plotted in Panel (a) of Figure (1) is quite noisy given that government statistics on incidences have been affected by changes in testing intensity and weekend reporting delays. These recording delays and corrections in the logging of cases introduce administrative noise. Therefore, to minimize noisy signals, we work with the 7-days moving average of the number of new cases depicted in Panel (a) of Figure (1).

Panel (b) of Figure (1) shows that Lombardy, with only a 16% of the total Italian population, was the epicenter of the pandemic in Italy during the first month of the outbreak, accounting approximately the 50% of the new and cumulative contagions. From May 2020 to the end of September 2020, the cumulative share of incidences with respect the country’s total, remained close to the 40% threshold. However, from September 2020 onwards, even if the incidence of Lombardy increased at a high path due to the unfolding of the second epidemic wave, other Italian regions where also highly affected, which explains the decrease in the Lombardy’s share.

**Figure 1:**
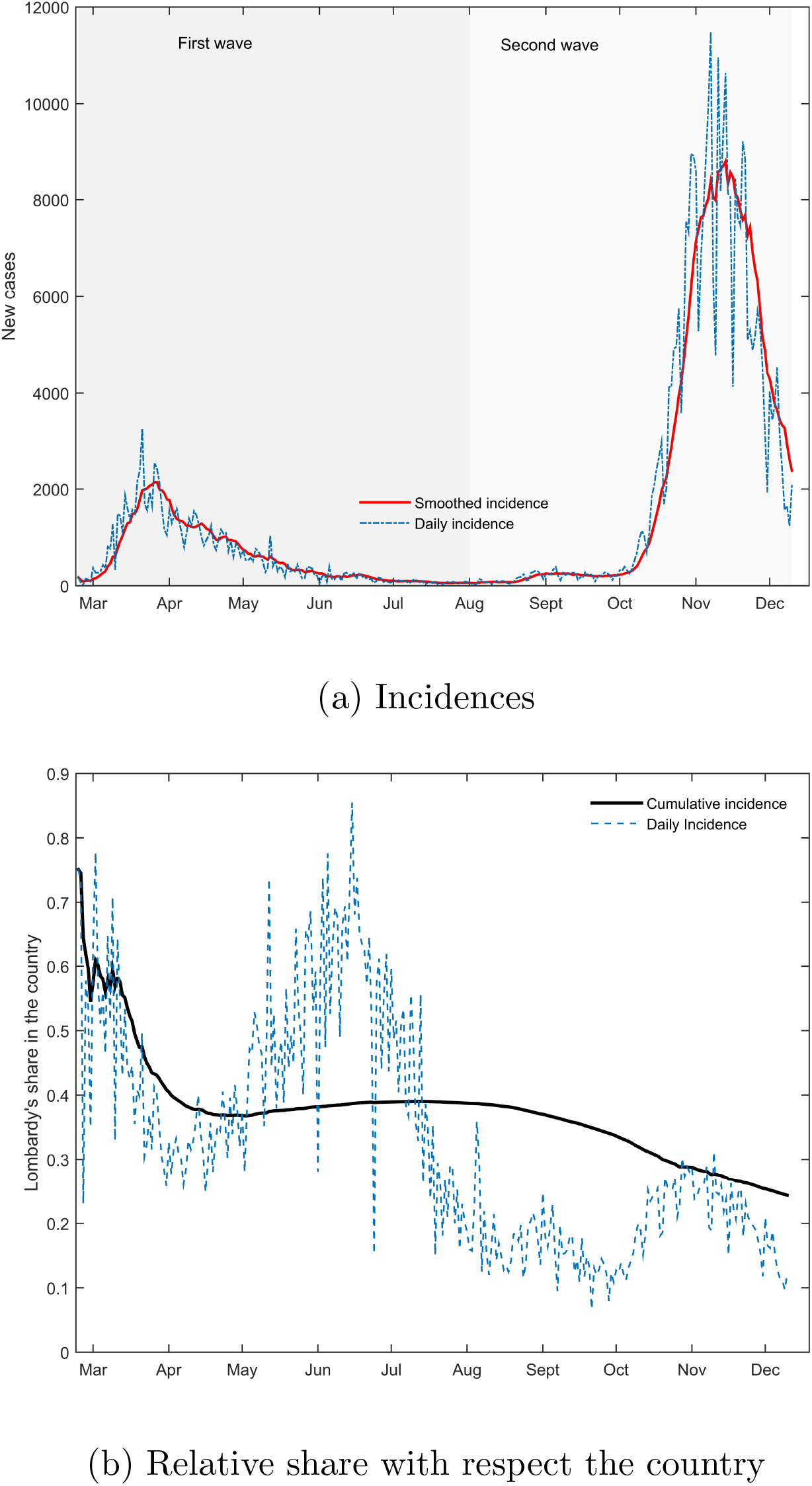
The dynamics of COVID-19 in Lombardy.

### 2.2 The predictors of the COVID-19 spread

There is no established forecasting model for the evolution of COVID-19 incidence. Thus, we now briefly review the literature analyzing the potential drivers of COVID-19 dynamics, and provide a brief justification for their consideration as driving forces behind the accelerations and slowdowns in the observed incidences. These factors capture variations in broader groups of determinants, namely: (i) epidemic dynamics, (ii) human mobility (iii) climatic conditions, (iv) environmental pollution and (v) health policy and containment measures. Table 1 presents the detailed definitions and sources of all the predictors used in the paper while additional information on the data set construction is included in the Appendix A.

**Table 1:**
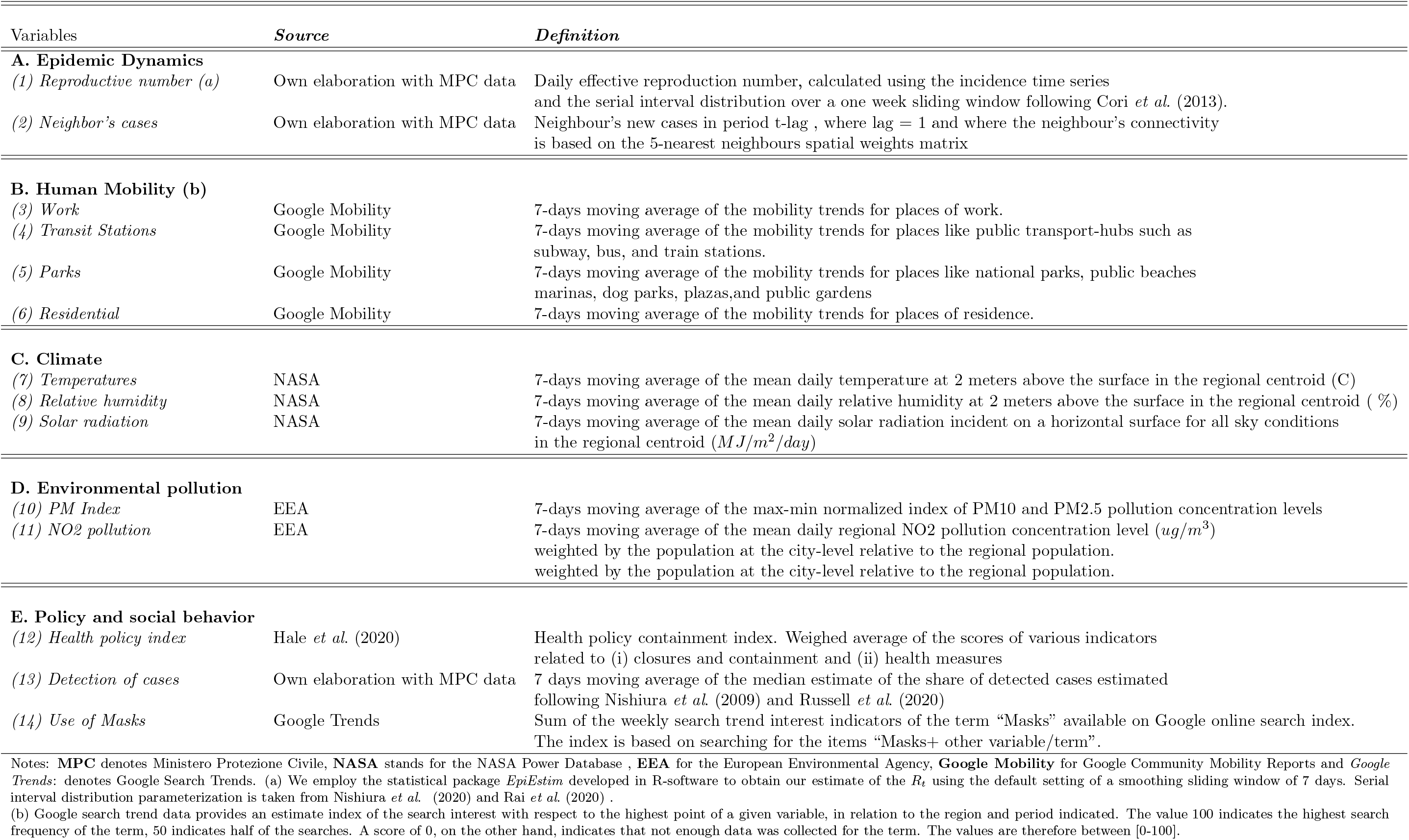
Data: Definitions and sources.

#### 2.2.1 Epidemic Dynamics

The first metric used to investigate the evolution of the COVID-19 incidence in Lombardy is the *(1) effective instantaneous reproductive number* (*R*_*t*_), which measures the average number of secondary cases per infectious case in a population made up of both susceptible and non-susceptible hosts (assuming that conditions remain identical after that time). In addition to the reproductive number, and in order to account for the possibility of importing cases from neighboring regions (Charaudeau *et al*., 2014; Andersen *et al*., 2020; Krisztin *et al*., 2020), we introduce *(2) the average incidence in neighboring regions*. This allows us to capture spillover/neighboring effects given that the mobility of infected individuals between regions may have contributed to the spread of the disease across borders.

### 2.2.2 Human Mobility

In the context of SARS-CoV2, which is propagated among people via small airborne micro-droplets (also commonly referred to as “aerosols”), larger respiratory droplets (which fall close to where they are expired) and direct contact with contaminated surfaces (fomites), a higher level human mobility reflects increased social interactions and possibilities of transmission (WHO, 2020). For this reason, higher citizen mobility has been shown to accelerate the diffusion of the virus among the population in a number of studies (see Ayyoubzadeh *et al*., 2020; Cartení et al., 2020;Kraemer *et al*., 2020; Chernozhukov *et al*., 2020; Zhou *et al*., 2020 among others). To exploit the link between individual mobility/ and the subsequent spread of the COVID-19 virus in our forecasting exercise, we rely on regional-level mobility data of Lombardy provided by the Google Mobility Reports. Specifically we employ mobility measurements on *(3) Workplaces, (4) Transit stations, (5) Residential areas* and *(6) Parks*.

#### 2.2.3 Climate

Disease agents and their vectors have specific environments that are optimal for growth, survival, transport, and dissemination, and climatic conditions define such environment (WHO, 2005; Makinen *et al*., 2009). The reason is that climate factors may affect not only the susceptibility conditions of the host by decreasing metabolic functions and defense barriers (Lowen and Steel, 2014), the physical properties of the virion envelope and its stability, but also the efficiency of the different routes of viral transmission (Duan *et al*., 2003; Van Doremalen *et al*., 2020). In the context of the COVID-19 epidemic, a large body of literature already exists suggesting epidemic curves are influenced by climate (see, Qi *et al*., 2020; Paez *et al*., 2020; Rios and Gianmoena, 2020). Using data from the NASA POWER v8 database we model climate effects by means of *(7) the mean temperature* 2 meters above the surface, the *(8) relative humidity* and the *level of ultra-violet (UV) solar radiation*.

#### 2.2.4 Pollution

Air pollution may exacerbate the vulnerability of populations to respiratory virus infections (Sciomer *et al*., 2020). As regards the evolution of the COVID-19 epidemic, different authors have hypothesized that another channel for a positive link between COVID-19 incidence and pollution is that airborne pollution particles may have been able to serve as carrier for the pathogen (Yongjian *et al*., 2020; Wu *et al*., 2020; Zoran *et al*., 2020). In fact, in the Italian context, there is evidence that suspended particulate matter pollution correlates positively with contagions and subsequent health damages (Fattorini and Regoli, 2020; Conticini *et al*., 2020). To use this potential source of predictability, we use daily air quality data taken from the European Environment Agency EEA. To measure the level of regional pollution, we employ a (10) Pollution Matter (PM) composite index aggregating PM10 and PM2.5 data. The other metric capturing the evolution of pollution is a *(11) NO2 pollution indicator*.

#### 2.2.5 Health Policy and Epidemic Monitoring

Health policy and containment measures, together with the ability to monitor its evolution are relevant to explain the evolution of COVID-19 (Deb *et al*., 2020; Chernozhukov *et al*., 2020). To model policy response effects we use a national level *(12) health policy containment composite index* developed by Hale *et al*. (2020) which contains information on a variety of closures, bans and restrictions. We also take into account the *(13) share of detected cases* with respect the total epidemic size in the region since the consensus of the literature is that the ability of regional authorities’ to perform tests and detect infections in real time is central in the strategy to curve the spread of the disease (Romer, 2020; Wang *et al*., 2020b).^5^ Finally, we employ Google Trends search data to target keywords related to the use of *(14) “face masks”* as previous studies of Lin *et al*. (2020) and Effenberger et al. (2020) find that online search data has predictive potential on the evolution of the epidemic and recent meta-analysis by Chu *et al*. (2020) and Liang *et al*. (2020) find the use of face-masks results in a large reduction in the risk of infection.

## 3 Econometric Methodology

### 3.1 Dynamic Model Averaging

Raftery *et al*. (2010) develop a method known as Dynamic Model Averaging (DMA). Later on, DMA and Dynamic Model Selection (DMS) approaches have been successfully employed in the field of macroeconomics (Koop and Korobilis, 2012), empirical finance (Naser, 2016, Drachal, 2016; Dong and Yoon, 2019), but also in the context of regional house price forecasting (Bork and Møller, 2015; Wei and Cao, 2017) by showing markedly improvements in forecasting accuracy with respect alternative modeling tools.

To see how DMA works, suppose that we have a set of *K* predictors. This implies a model space of size 2^*K*^ models, made by different combinations of these *K* predictors. Denoting these models *M*_*k*_ for *k* = 1, …, 2^*K*^ by the specific subset/combination of regressors *X*^(*k*)^, our set of models can be written as:

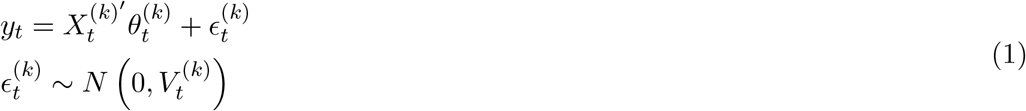

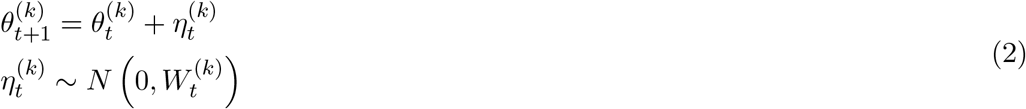

where *y*_*t*_ is the dependent variable to be forecasted. As explained before, in the context of this study, *y*_*t*_ are smoothed data on the daily new cases. *X*_*t*_ is a 1×*K* vector of predictors for our dependent variables, *θ*_*t*_ is a *K* × 1 vector of coefficients (states), ϵ_*t*_ ∼ *N* (0, *V*_*t*_) and *η*_*t*_ ∼ *N* (0, *W*_*t*_). The errors ϵ_*t*_ and *η*_*t*_ are assumed to be mutually independent at all leads and lags. Therefore, no systematic movement in the time-varying parameters is assumed and the changes in *θ*_*t*_ are unpredictable a priori.

In the DMA framework, Equation (1) is labeled the **measurement equation**, whereas Equation (2) receives the name of **transition** or **state equation**. The measurement equation allows the parameters to be time-dependent while the transition equation determines the movement of the parameters. The conditional variances *V*_*t*_ and *W*_*t*_ are unknown quantities associated with the measurement equation and the states equation.^6^

Let *L*_*t*_ ∈ 1, 2, …, 2^*K*^ denote which model *M*_*k*_ applies at each time period, 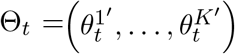 and *Y* ^*t*^ = (*y*_1_, …, *y*_*t*_) ′. Thus, if *L*_*t*_ = *k*, the process is governed by model *M*_*k*_ at time *t*. The fact that different models hold at each time, and we will do model averaging, justifies the terminology “dynamic model averaging”. When forecasting time *t* variables using information through time *t*−1, DMA involves calculating the probability *Pr L*_*t*_ = *k*|*Y* ^*t*−1^ for *k* = 1, …, 2^*K*^ and averaging forecasts across the 2^*K*^ potential models formed by combinations of predictors using these probabilities.

In this setting, the evolution of models over time can be determined by a 2^*K*^ × 2^*K*^ transition probability matrix, *P*, determining how predictors enter/leave the model with elements *p*_*k,l*_ = *Pr* (*L*_*t*_ = *k*|*L*_*t*−1_ = *l*) for *k, l* = 1, …, 2^*K*^. Nevertheless, unless the number of predictors *K* is very small the transition probability matrix *P* will have so many entries that inference will be imprecise and computation slow, rendering a full Bayesian approach to DMA quite difficult (Koop and Korobilis, 2012). Thus, the fundamental challenge of the modeling approach given by Equations (1) and (2) is how to compute the evolution of models over time. To achieve a feasible computation, we follow Raftery *et al*. (2010) and Koop and Korobilis (2012), who propose an approach that involve two forgetting factors *α* and *λ*, which are fixed to numbers slightly below one and help to produce an evolution of parameter estimates and model probabilities based on age-weighted data.

In our model setup the underlying state is characterized by the pair (Θ_*t*_, *L*_*t*_) and the probability distribution of (Θ_*t*_, *L*_*t*_) is given by:

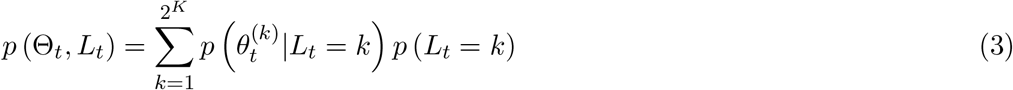

which will be updated each time as new data becomes available. The estimation of our state space multi-model framework uses an adaptive strategy based on the Kalman filter and consists of a *prediction* and an *updating* step for both, parameters and models. We begin by describing (i) the prediction and updating steps of the parameters and then we move to the one of (ii) the models.

Specifically, the Kalman filtering estimation begins with the result that:

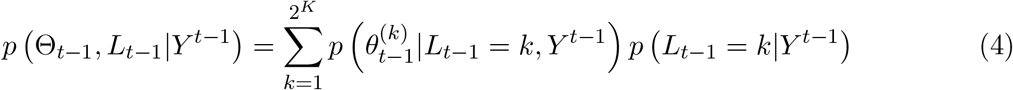

where 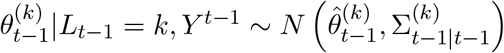

Then, the filter proceeds by predicting 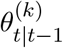 using all information available up to time *t* − 1, (*Y* ^*t*−1^ = *y*_1_, …, *y*_*t*−1_) as:

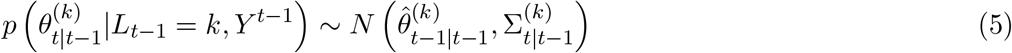

where the variace-covariance matrix of the states at period *t* conditional on the information at *t* − 1 is:

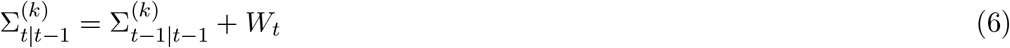

Raftery *et al*. (2010) propose to avoid the estimation or simulation of *W*_*t*_ and simplify Equation (6) by using:

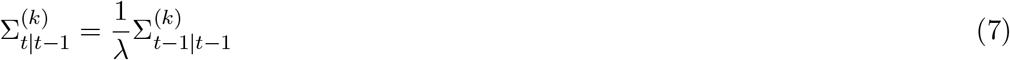

where 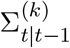 denotes the covariance matrix of 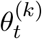 and 0 ≤ *λ* ≤ 1. ^7^ The value of the forgetting factor *λ* determines how rapidly the parameters of the model evolve (i.e, a high value of *λ* implies a higher stability, whereas a low value of *λ* produces rapid changes in the parameters).^8^

In a second step, the parameters are updated as follows:

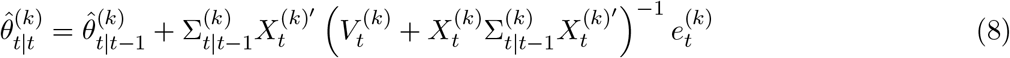

where 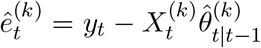 is the 1-period step ahead forecast error and the variancecovariance matrix of 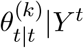 evolves as:

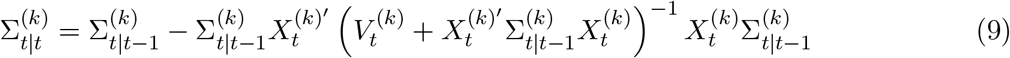

The term 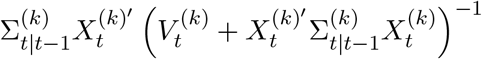 is usually called the “Kalman gain”, which minimizes the posterior error covariance and is informative on how much correction we should take from measurements *y*_*t*_ when updating the states, *θt*|*t*. High values of the Kalman gain make the filter more responsive to recent measurements whereas in the case of low values of the Kalman gain, the filter follows the state predictions more closely decreasing the variability of *θ*_*t*_ over time.

Recall that to achieve a computationally feasible estimation of the time-varying parameters and avoid the burdensome Markov Chain Monte Carlo (MCMC), we introduced *λ* to prevent the estimation of 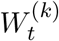. We now proceed similarly for the model probabilities by introducing a forgetting factor *α*. The model prediction equation using the Kalman filter is illustrated by:

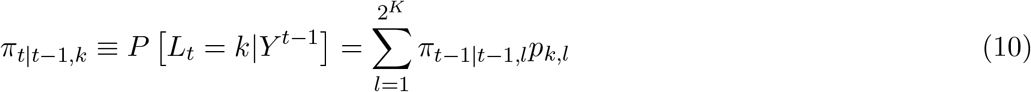

However, as mentioned above, instead of specifying the transition probability matrix *P* we use an approximation following Raftery *et al*. (2010) that replaces Equation (10) by:

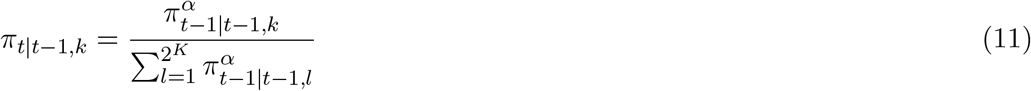

where *α* is the model probability forgetting factor, 0 *< α* ≤ 1, and it is interpreted in a similar manner to *λ*. Equation (11) implies that if a specific combination of regressors forming *M*_*k*_ forecasts well in the recent past, it will received more weight at time *t*. However, the lower the value of *α* the lower the weight is given to models that performed well forecasting during the past relative to models with good forecast performance last period.^9^ As noted by Koop and Korobilis (2012), the main advantage of using *α* instead of drawing transitions between models is that it greatly simplifies the computational burden of the exercise since we only need *πt*|*t*−1,*k* and *πt*−1|*t*−1,*k* to proceed.

Finally, the model updating equation is defined as:

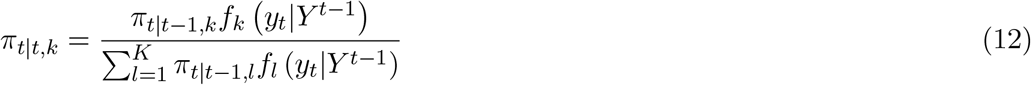

where *f*_*l*_ (*y*_*t*_|*Y* ^*t*−1^) is the predictive density for model *l* evaluated at *y*_*t*_ given by:

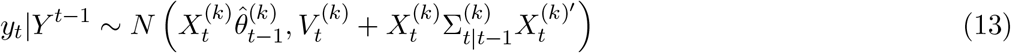

Conditional on 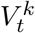 the estimation strategy discussed above only involves evaluating previous formulae given an initializing prior for *π*_0,0,*k*_ and 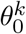 for *k* = 1, …, 2^*K*^. While Raftery *et al*. (2010) reccommends using a plug in method where 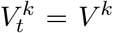 Koop and Korobilis (2012) recommend to account for the possibility that the error variance is changing over time. We follow Koop and Korobilis (2012) and assume 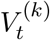 can be modeled by an Exponentially Weighted Moving Average (EWMA):

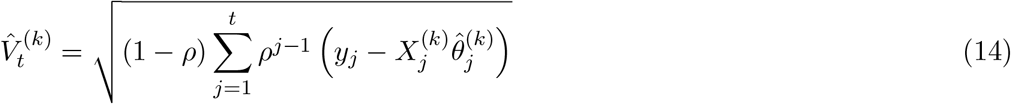

where *ρ* is a decay factor, and the period *t* + 1 forecast given data up to time *t* takes the form of the following recursion:

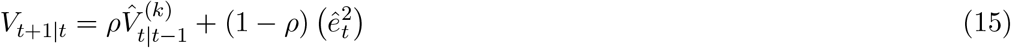

where we set the value of *ρ* to 0.95.

Finally, note that recursive forecasting of the dependent variable in the DMA can be done at each point in time by taking the probabilistic weighted average of all possible models according to the probabilities *πt*|*t*−1,*k* :

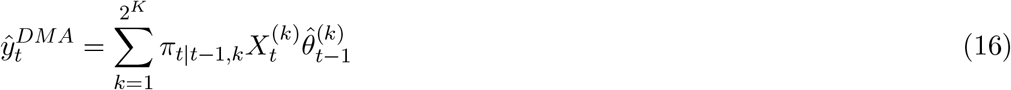

The difference of DMS with DMA is that DMS proceeds by selecting the single model with the highest value for 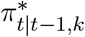 at each point in time, and simply using it for forecasting. Hence, the forecast implied by the DMS procedure is given by:

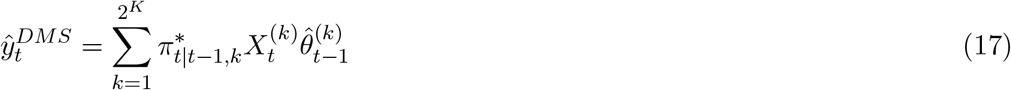

### 3.2 Model Evaluation

There are many metrics for evaluating the forecast performance of a model, but the majority of research in epidemic forecasting pays attention to producing and evaluating point forecasts (see Roosa *et al*., 2020a,b; Roda *et al*., 2020; Chowell *et al*., 2020).

Point forecasts receive this high attention in the forecast evaluation process because they are easy to compute and understand. However, focusing on point forecasts alone in the context of epidemics has been criticized by Ioannidis *et al*. (2020) and Taleb *et* al. (2020) on different grounds. Because of the evolution of epidemic curves is subject to a very high uncertainty, we will employ not only point forecasts but also interval and density forecasts to derive a variety of loss functions as evaluation criteria of the DMA/DMS approaches.

The standard Bayesian metric for density forecast comparison is the Average of the sum of Log Predictive Likelihoods (ALPL) (Geweke and Amisano, 2011), which involves the entire predictive distribution and not simply point forecasts. The predictive likelihood is the predictive density for *y*_*t*_ given data through time *t* − 1 evaluated at the actual outcome (i.e, in model *M*_*k*_, the predictive density is *p*_*k*_ *y*_*t*_|*Y* ^*t*−1^).^10^

In addition to the ALPL we also consider the Mean Absolute Percentage Forecast Error (MAPFE) which is defined as:

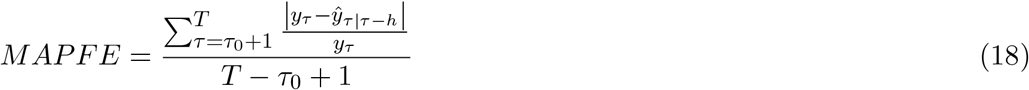

where *ŷ*_*τ |τ −h*_ is the point forecast of *y*_*τ*_ using the information available at time *τ* – *h* where *h* is the forecast horizon.^11^.

We also compute 95% confidence interval coverage rates for each model (i.e.,the percentage of times in which the actual number of contagions is contained in the forecast confidence interval) as an accurate assessment of the uncertainty surrounding forecasts is likely to be of interest for health authorities and policy-makers. A model that delivers coverage rates which are very low when compared to alternative competing models would underestimate forecast uncertainty. Therefore, we calculate the (iii) the 95%Prediction

Interval Coverage (PIC) as:

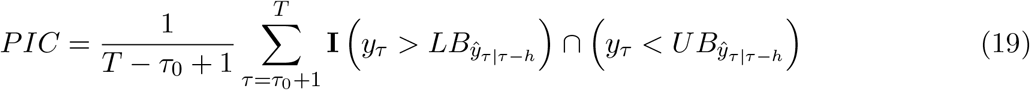

where 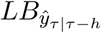 and 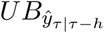 are the lower and upper bounds of the 95% prediction intervals respectively and **I** is an indicator variable that equals 1 if *y*_*τ*_ is in the specified interval, and 0 otherwise. An issue with the PIC is that in the extreme, coverage rates of a 100% could imply that the estimated forecast confidence intervals always contain the actual values, but this could be at the cost of the confidence bands being so wide that are of little practical use. Hence, to complement this metric, we rely on (iv) the Mean Interval Score (MIS) proposed by Gneiting and Raftery (2007), which in the field of epidemiology has also been used by Chowell *et al*. (2020). The MIS considers the width of the interval as well as the coverage, with a penalty for data points not included within the prediction intervals. Therefore, the forecaster is rewarded for narrow prediction intervals, and he or she incurs a penalty, the size of which depends on the significance level, if the observation misses the interval. For a significance level of the 5%, the MIS is calculated as:

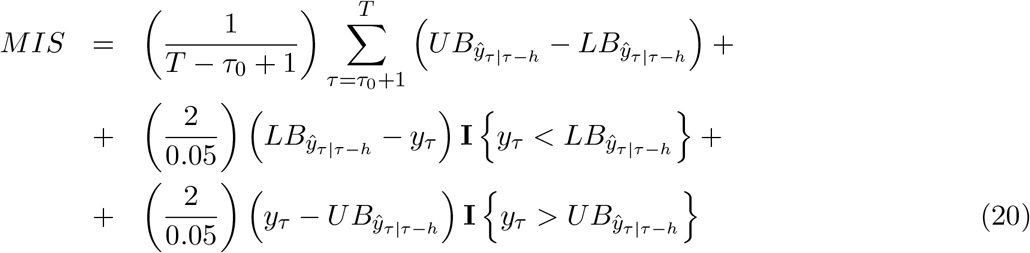

## 4 Results

We now turn our attention to our results, which are divided in two subsections.

The first subsection investigates forecast performance by comparing DMA and DMS forecasts with those produced by several alternative competing strategies by looking at the different aforementioned metrics evaluated at different horizons. As it is common in the literature we consider short term *h* = 1, 3, 7 and 14 step-ahead daily forecasts.

The second of these subsections presents evidence on which variables are good for predicting COVID-19 contagions over the time-sample considered. It presents the results using DMA, implemented with three different configuration for forgetting factors which involve setting (i) *α* = *λ* = 0.99, (ii) *α* = *λ* = 0.95 and (i) *α* = *λ* = 0.90, a noninformative prior over the models (i.e, 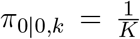 for *k* = 1, …, *K* so that initially all models are equally likely) and a diffuse prior on the initial conditions of the states: 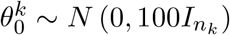 where *n*_*k*_ is the number of variables in model *k*.

### 4.1 Forecasting Models and Forecast Performance Analysis

We begin to forecast COVID-19 daily incidence by using AR(1)-X type models of the form:

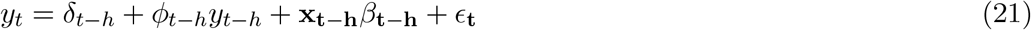

where *y*_*t*+*h*_ denotes the *h*-steps ahead daily COVID-19 incidence regressed on an intercept, a time lag and exogenous predictors. We implement direct forecasts for *h >* 1 for practical reasons given that iterated forecasts would require predictive simulation which in the context of a model space of the magnitude considered here would be computationally burdensome. ^12^

We implement our forecasts recursively that is, using an expanding window so that all available information at the time of the forecast is used to estimate the models. We begin forecasting the first of March 2020, and use the periods of the 24 of February to the 29 of February as our first estimation period to forecast *h*-steps ahead. We then add one new observation to the estimation sample and forecast *h*-steps ahead until the full sample is exhausted. We use a variety of forecasting models. Note that some of the models employed in this exercise are based on Equation (21) but assume constant coefficients (i.e, *α*_*t*−*h*_ = *α, φ*_*t*−*h*_ = *φ* and so on). We now briefly describe our list of models (for further details see Appendix B):

### Dynamic Model-Averaging and Model-Selection

- DMA: Dynamic Model Averaging. Uses model probabilities as weights to compute the average forecast as in Raftery *et al*. (2010) and Koop and Korobilis (2012). We employ three different configurations, by setting the forgetting factors to (i) *α* = *λ* = 0.99, (ii) *α* = *λ* = 0.95 and (iii) *α* = *λ* = 0.90. We set *κ* = 0.95 in the three configurations.
- DMS: Dynamic Model Selection. Puts all the weight on the model with the highest probability to compute the forecast as Raftery *et al*. (2010) and Koop and Korobilis (2012). We employ three different configurations by setting the forgetting factors to (i) *α* = *λ* = 0.99 (ii) *α* = *λ* = 0.95 and (iii) *α* = *λ* = 0.90. We set *ρ* = 0.5 in the three configurations.

### Time-series and Machine Learning

- TVP-AR(1): Time-varying parameter AR(1) model, including only an intercept and a time lag (without any of the predictors), estimated with the Kalman filter using as forgetting factors *λ* = *ρ* = 0.95.
- TVP-AR(1)X: Time-varying parameter AR(1) model with intercept, a time lag and the full set of *X* regressors, estimated with the Kalman filter using as forgetting factors *λ* = *ρ* = 0.95
- TVP-SV-AR(1): Time-varying parameter AR(1) model with stochastic volatility, including an intercept and a time lag (without any of the predictors) estimated with the MCMC algorithm of Chan and Jeliazkov (2009).^13^
- TVP-SV-AR(1)X: Time-varying parameter AR(1) model with stochastic volatility, including an intercept, a time lag and the full set of *X* regressors estimated with the MCMC algorithm of Chan and Jeliazkov (2009)
- BMA: Bayesian Model Averaging. DMA with forgetting factors fixed at *α*_*t*_ = *λ*_*t*_ = *ρ* = 1)
- BMS: Bayesian Model Selection. DMS with forgetting factors fixed at *α*_*t*_ = *λ*_*t*_ = *ρ* = 1)
- BSSVS: Bayesian Stochastic Variable Search Selection. Builds on benchmark AR(1)X specification, estimated using the SSVS prior with MCMC of George and McCulloch (1993).
- BAG: Bagging. Same predictors as TVP-AR(1)X, estimated as constant parameter regression using the Bagging algorithm of Breiman (1996).
- PLS: Partial Least Squares. Same predictors as TVP-AR(1)X, estimated as a constant parameter Partial Least Squares regression using the SIMPLS algorithm of DeJong (1993) and retaining *K* factors.

### Epidemic mathematical models

In addition to these time-series and machine learning approaches, we also employ modern modeling approaches widely employed in the field of epidemics. Specifically, we consider models of epidemic growth, based on phenomenological and compartmental approaches.

Phenomenological considered here models have in common the assumption of a decay in the growth rate of the epidemic as the total number of contagions increases. A nice property of these models is that they provide a good model for the exponential growth phase and include the so called saturation mechanism leading to the equilibrium, with a cumulative number of contagions stabilization after some point in time. The set of phenomenological models is estimated with quantified uncertainty following Chowell (2017), Burger *et al*. (2019) and Roosa *et al.* (2020a,b).^14^

- GLGM: Generalized Logistic Growth Model. The model is given by:

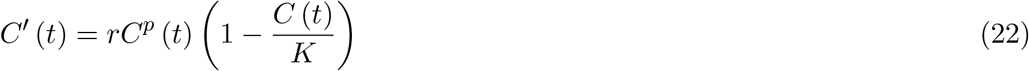

where *C* (*t*) is the cumulative cases at time *t, C* ′ (*t*) is the daily incidence, *r* is the growth rate and *K* is the carrying capacity. The parameter *p* ∈ [0, 1] is a scaling of growth factor, that accommodates sub-exponential growth patterns in the spread of the disease (see)

- GRGM: Generalized Richards Growth Model. The model is given by:

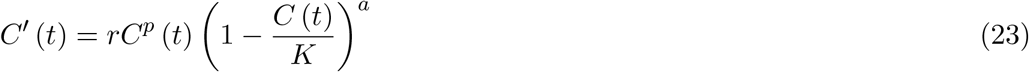

where *a* is a parameter used to capture the deviation of the symmetric S-shaped dynamics of the simple logistic growth model.

- Susceptible-Infected-Removed (SIR) model. The SIR model classifies individuals in the compartment as one of three classes: susceptible (S), infectious (I), and recovered or removed (R). Infectious individuals spread the disease to susceptible individuals at rate *β* and remain in the infectious class for a given period of time known as the infectious period before moving into the recovered (or removed) class at rate *γ*. Individuals in the recovered class are assumed to be immune for an extended period (or removed from the population). For the total population *N* = *S* + *I* + *R*, the dynamical system describing the SIR equations is given as:

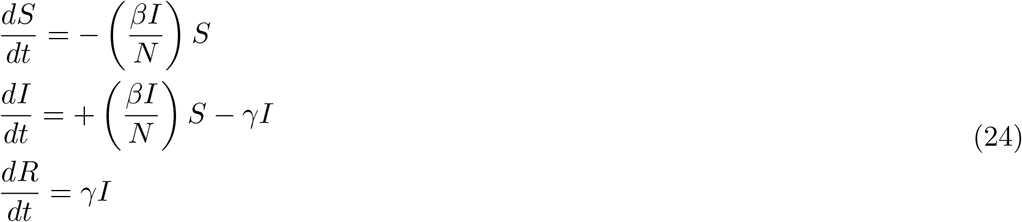

Given the initial conditions S(0), I(0) and R(0), we estimate the classical SIR model by means of Maximum Likelihood techniques assuming a Poisson distribution. To connect the model with the data, we will use the following measurement equation: 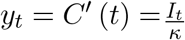, where 1*/κ* is a combination of the reporting rate, the asymptomatic rate, and the total population size.

The choice of models is based on their simplicity, replicability and popularity. Forecast accuracy results are measured using the (i) MAPFE, (ii) PIC, (iii) MIS and (iv) ALPL metrics described in Section (3.2) and presented in Tables (2) to (4). The main story coming out of these tables is clear: DMA and DMS forecast better than the other approaches in terms of density and interval forecasts at short horizons, and never much worse than the best alternative as regards to point forecasts.

**Table 2:** Forecasting results: Density Forecasts.

| Average Log Predictive Likelihoods (ALPL) |  |  |  |  |
| --- | --- | --- | --- | --- |
| | $h = 1$ | $h = 3$ | $h = 7$ | $h = 14$ |
| TVP-AR(1) | -5.92 | -8.64 | -13.29 | -12.26 |
| TVP-AR(1)X | -5.95 | -8.73 | -13.61 | -14.02 |
| TVP-AR(1)-SV (MCMC) | -9.25 | -16.16 | -18.90 | -17.14 |
| TVP-AR(1)X-SV (MCMC) | -9.09 | -8.64 | -11.96 | -13.66 |
| TVP-DMA ($\alpha = \lambda = 0.99$) | -5.95 | -8.38 | -13.13 | -14.78 |
| TVP-DMA ($\alpha = \lambda = 0.95$) | -5.94 | -8.89 | -12.13 | -15.70 |
| TVP-DMA ($\alpha = \lambda = 0.90$) | **-5.63** | **-8.35** | -13.41 | -15.40 |
| TVP-DMS ($\alpha = \lambda = 0.99$) | -8.12 | -10.18 | **-11.03** | **-11.39** |
| TVP-DMS ($\alpha = \lambda = 0.95$) | -8.09 | -10.62 | -11.60 | -12.94 |
| TVP-DMS ($\alpha = \lambda = 0.90$) | -7.79 | -10.24 | -11.93 | -14.41 |
| BMA | -12.33 | -19.78 | -28.14 | -31.30 |
| BMS | -15.14 | -19.94 | -24.19 | -27.39 |
| SSVS | -7.37 | -10.54 | -15.02 | -18.00 |
| PLS | -7.88 | -11.05 | -16.83 | -20.38 |
| BAG | -8.68 | -11.82 | -16.71 | -21.00 |
| GLGM | -16.45 | -15.97 | -16.74 | -16.14 |
| GRGM | -17.99 | -18.37 | -18.68 | -18.70 |
| SIR-ML | -14.41 | -14.75 | -14.58 | -15.54 |

Notes: Entries in columns 2-5 of this Table are mean represent the Average Log Predictive Likelihood. Higher values of the ALPL signify a deterioration of the quality of the forecast whereas lower values signify an improvement. Entries in boldface indicate the best performing model for each forecast statistic and for each forecast horizon.

**Table 3:** Forecasting results: Point Forecasts.

| Mean Absolute Percentage Forecast Error (MAPFE) |  |  |  |  |
| --- | --- | --- | --- | --- |
| | $h = 1$ | $h = 3$ | $h = 7$ | $h = 14$ |
| TVP-AR(1) | 0.063 | 0.212 | 0.841 | 0.725 |
| TVP-AR(1)X | 0.071 | 0.232 | 0.583 | 0.866 |
| TVP-AR(1)-SV (MCMC) | **0.052** | **0.100** | **0.305** | 0.847 |
| TVP-AR(1)X-SV (MCMC) | 0.050 | 0.097 | 0.265 | 0.731 |
| TVP-DMA ($\alpha = \lambda = 0.99$) | 0.069 | 0.220 | 0.571 | 2.516 |
| TVP-DMA ($\alpha = \lambda = 0.95$) | 0.057 | 0.161 | 0.422 | 1.169 |
| TVP-DMA ($\alpha = \lambda = 0.90$) | 0.058 | 0.133 | 0.380 | 0.793 |
| TVP-DMS ($\alpha = \lambda = 0.99$) | 0.099 | 0.254 | 0.520 | 1.877 |
| TVP-DMS ($\alpha = \lambda = 0.95$) | 0.082 | 0.199 | 0.358 | 0.628 |
| TVP-DMS ($\alpha = \lambda = 0.90$) | 0.081 | 0.170 | 0.331 | 0.554 |
| BMA | 0.078 | 0.172 | 0.511 | 1.052 |
| BMS | 0.105 | 0.199 | 0.382 | 0.659 |
| SSVS | 0.094 | 0.299 | 0.740 | 1.353 |
| PLS | 0.081 | 0.263 | 0.904 | 1.535 |
| BAG | 0.080 | 0.252 | 0.836 | 1.526 |
| GLGM | 0.372 | 0.410 | 0.448 | 0.528 |
| GRGM | 0.393 | 0.449 | 0.499 | 0.590 |
| SIR-ML | 0.269 | 0.300 | 0.366 | **0.510** |

Notes: Entries in columns 2-5 of this Table are mean represent the Mean Absolute Percentage Forecast Errors. Higher MAPFE values signify a deterioration of the quality of the forecast whereas lower values signify an improvement. Entries in boldface indicate the best performing model for each forecast statistic and for each forecast horizon.

**Table 4:** Forecasting results: Interval Forecasts.

|  | 95% PIC |  |  |  | MIS |  |  |  |
| --- | --- | --- | --- | --- | --- | --- | --- | --- |
| | $h = 1$ | $h = 3$ | $h = 7$ | $h = 14$ | $h = 1$ | $h = 3$ | $h = 7$ | $h = 14$ |
| TVP-AR(1) | 0.897 | 0.773 | **0.737** | **0.645** | 883.3 | 4,252.7 | 24,160.7 | 34,476.5 |
| TVP-AR(1)X | 0.915 | 0.770 | 0.626 | 0.676 | 869.4 | 5,432.8 | 22,435.3 | 46,664.3 |
| TVP-AR(1)-SV (MCMC) | 0.688 | 0.363 | 0.241 | 0.309 | 1,217.7 | 3,357.6 | 14,991.9 | 42,242.9 |
| TVP-AR(1)X-SV(MCMC) | 0.589 | 0.615 | 0.581 | 0.449 | 1,330.3 | 2,803.7 | 12,477.3 | 48,689.1 |
| TVP-DMA ($\alpha = \lambda = 0.99$) | 0.911 | 0.770 | 0.652 | 0.469 | 823.8 | **2,720.1** | 15,971.0 | 71,086.4 |
| TVP-DMA ($\alpha = \lambda = 0.95$) | 0.911 | 0.763 | 0.670 | 0.508 | 796.9 | 3,992.3 | 14,417.0 | 58,442.6 |
| TVP-DMA ($\alpha = \lambda = 0.90$) | **0.935** | **0.798** | 0.670 | 0.559 | **628.1** | 3,013.3 | 20,115.6 | 55,768.2 |
| TVP-DMS ($\alpha = \lambda = 0.99$) | 0.752 | 0.680 | 0.656 | 0.598 | 1,601.3 | 4,918.5 | 11,006.6 | 25,704.3 |
| TVP-DMS ($\alpha = \lambda = 0.95$) | 0.780 | 0.698 | 0.674 | 0.578 | 1,386.5 | 4,703.8 | **10,292.0** | 26,172.1 |
| TVP-DMS ($\alpha = \lambda = 0.90$) | 0.798 | 0.727 | 0.670 | 0.563 | 1,279.3 | 4,168.4 | 10,795.7 | **25,094.8** |
| BMA | 0.635 | 0.360 | 0.148 | 0.082 | 1,951.9 | 7,406.7 | 26,864.7 | 60,539.0 |
| BMS | 0.543 | 0.371 | 0.259 | 0.184 | 3,348.2 | 10,422.7 | 27,913.4 | 48,423.4 |
| SSVS | 0.801 | 0.701 | 0.563 | 0.461 | 1,126.1 | 4,503.5 | 20,156.2 | 59,870.6 |
| PLS | 0.780 | 0.640 | 0.404 | 0.336 | 1,114.9 | 4,342.9 | 22,336.3 | 54,716.2 |
| BAG | 0.759 | 0.665 | 0.430 | 0.340 | 1,284.9 | 5,604.2 | 23,020.9 | 55,295.4 |
| GLGM | 0.164 | 0.153 | 0.133 | 0.113 | 29,209.4 | 32,278.0 | 38,140.0 | 46,884.9 |
| GRGM | 0.105 | 0.097 | 0.079 | 0.063 | 31,770.4 | 36,259.5 | 44,863.5 | 55,407.2 |
| SIR - ML | 0.099 | 0.091 | 0.086 | 0.083 | 22,166.53 | 27,334.29 | 39,491.74 | 64,629.79 |

Notes: Entries in columns 2-5 of this Table are mean 95% Prediction Interval Coverage (PIC) rates, and columns 6-9 are average Mean Interval Scores (MIS). Entries for each model are PICR and MIS values. Higher PICR scores signify improvement whereas lower values signify a deterioration. Lower MIS values signify an improvement whereas higher values a deterioration. Entries in boldface indicate the best performing model for each forecast statistic and for each forecast horizon.

Considering first log predictive likelihoods shown in Table (2), which is the preferred method of Bayesian forecast comparison, we find that DMA or DMS forecast best, than the other forecasting strategies used in our comparison exercise at *h* = 1 and *h* = 14. Note the excellent performance of DMA *α* = *λ* = 0.90 for short run horizons *h* = 1 and *h* = 3. This value for the forgetting factors allows for rapid change in both coefficient and in models. Versions of DMA that impose more gradual change do slightly worse, but DMS versions with slower model and parameter change (i.e, *α* = *λ* = 0.99) obtain the highest ALPLs for longer horizons *h* = 7 and *h* = 14. As regards the MAPFE, which is our point forecast performance metric, the results of Table (3) show that the DMA for *h* = 1 and *h* = 3 tend to outperform the DMS, but that the DMS can achieve one of the lowest percentage errors at *h* = 14 steps ahead. In both metrics we find strong evidence that allowing for faster model and parameter variation tends to increase accuracy. This is evident when comparing the three DMA configurations given that as we decrease *α* and *λ* the ALPL increases and the MAPFE of the DMA decreases.

Regarding the results of the MAPFE, we find that DMA/DMS are weakly dominated by the TVP-AR(1)X-SV estimated by means of MCMC and with stochastic volatility, at least for short run and medium horizons. The MAPFE of the TVP-AR(1)X-SV are the 5%, 9.7% and 26.5% for *h* = 1, 3, and 7 whereas the lowest DMA/DMS MAPFEs for these horizons are 5.7%, 13.3% and 33% respectively. However, for *h* = 14 the errors of DMS with *α* = *λ* = 0.9 (55.4%), are much lower than those of the TVP-SV-AR(1)X (84.7%). Taken together these results suggest that the optimal forecasting strategy would be a DMA/DMS with stochastic volatility estimated with MCMC rather than with forgetting factors but that would render the estimation too slow. Phenomenological models perform poorly in the short run irrespective but appear to be close in accuracy to the DMA/DMS for *h* = 14. This is also the case of the SIR model as it produces the lowest error at *h* = 14.

PIC and MIS results reveal that DMA/DMS are the best candidates when considering the uncertainty in the forecast for short run horizons. For *h* = 1 step ahead forecasts the coverage of DMA (*α* = *λ* = 0.9) is the 93.5% and for *h* = 3 it is the 78.8%. Interestingly, for horizons *h* = 7 and *h* = 14 the TVP-AR(1) and TVP-AR(1)X, estimated with forgetting factors, produce a higher coverage of the 73.7% and 67.6% respectively. However as revealed by the MIS, this higher coverage rate comes at the cost of producing too wide confidence bands, as it is clear the MIS for longer forecast horizons for these models is lower. In fact, for the MIS metric, the different DMA/DMS configurations outperform the other approaches.

Taken together, the results of our forecasting exercise suggest that both model change and parameter change help to improve accuracy. This can be seen in (i) the superior relative performance of TVP-AR(1)X with respect static parameter modeling approaches such as the SSVS, the PLS or the Bagging and in (ii) the performance of the TVP-DMA/DMS with respect the BMA and BMS.

As refers the importance of the information contained in exogenous predictors, the evidence is more mixed. In terms of point and density forecasts we find that TVP-SV-AR(1)X outperforms the TVP-SV-AR(1) for *h* = 1, 3 and *h* = 7, but the TVP-AR(1) model does the same with respect the TVP-AR(1)X. Regarding the production of accurate confidence bands, as measured by the PIC and the MIS, the specifications including the information of exogenous predictors tend to dominate for *h* = 7 and *h* = 14 but not for *h* = 1. In any case, DMA and DMS produce an automatically selected degree of shrinkage which in all cases leads to superior forecasts when compared to the extreme cases of the TVP-AR(1)X and the TVP-AR(1).

Finally, what do Tables (2) to (4) say about the relative forecast performance of DMA and DMS?. In this regard, what we find is that DMA with (*α* = *λ* = 0.90) seems better suited than DMS for short run forecasts whereas DMS (*α* = *λ* = 0.99) does a better job when producing long run forecasts.

### 4.2 Which variables are good predictors for COVID-19 spread?

Off the different forecasting approaches in the preceding section only DMA and DMS allow for different forecasting models at different times. Accordingly, in this section we focus only in these two approaches. Given the huge number of models explored at each time (i.e, 2^14^ = 16, 384), we cannot possibly present empirical results for every model. Instead, we summarize our results in two different ways. We begin with Figure (2) which illustrates that, although we have fourteen predictors which could be selected to forecast daily COVID-19 incidence, most of the time DMA attaches the highest probability to parsimonious models including only a few predictors.

**Figure 2:**
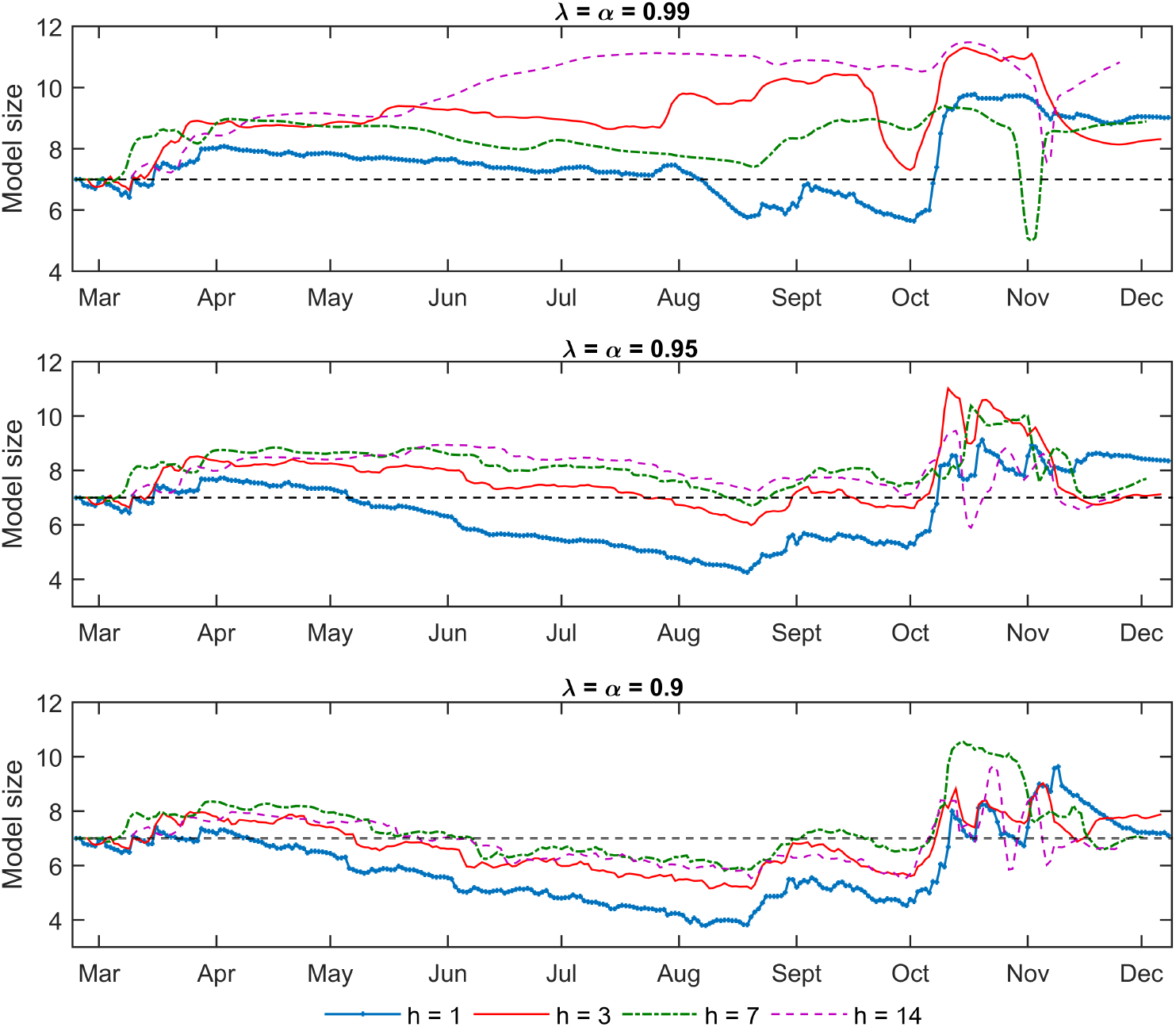
Expected model size.

If we let *Size*_*k,t*_ be the number of predictors in model *M*_*k*_ at time *t*, then we can calculate the number of expected predictors included in a DMA at time *t* as:

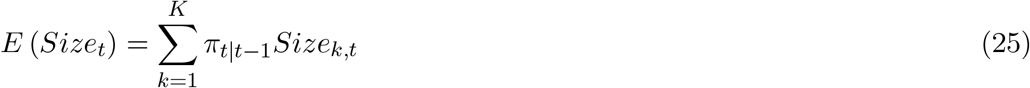

Figure (2) plots the value of *E* (*Size*_*t*_) for *h* = 1, 3, 7 and 14 days ahead for the various configurations of the forgetting factors. This figure gives an indication of variations in the degree of parsimony over time and across DMA configurations. As observed, Figure (2) shows that the number of predictors used by DMA changes over time and that the higher the degree of parameter variation, the higher the shrinkage. Another key feature of our empirical results is that, although we have 14 potential predictors (excluding the constant and the autoregressive term which are common to all models), most probability is attached to parsimonious models including few predictors. This result holds irrespective of the forecast horizon. Nonetheless, as it frequently occurs in DMA analysis, at longer forecast horizons of *h* = 14 days ahead, slightly more predictors are included in the forecasting model specification.

The pattern shown in Figure (2) indicates that forgetting factor configurations generate different model sizes with different variables inside them, with different estimates of variable importance. For *λ* = *α* = 0.99 as time goes by, and more data is available for estimation, more predictors are chosen. However, it can be observed that the period that ranges from October 2020 to December 2020 is characterized by abrupt changes in the model size (see the the abrupt drop in the number of predictors for medium to long term forecast horizons *h* = 7, 14 during November 2020). On the other hand, for the forgetting factor configurations *λ* = *α* = 0.95 = 0.90 which allow a higher degree of parameter and model variability, we find a different pattern. In these configurations the expected model size falls until mid-August 2020 and raises substantially during the second wave.

Taken together, these findings suggest that (i) when forecasting at longer horizons, using additional exogenous information to that of the past state of the epidemic tends to improve forecast performance. A second issue is that (ii) DMA results are quite sensitive to the forgetting factors and that the optimal model size when forecasting at shorter horizons is quite different to that of longer ones. This second point, together with the fact that we are performing a reduced form forecasting exercise, precludes us to provide too many stories on specific variables’ results or any interpretation on causal effects.

We now use the posterior inclusion probabilities (PIPs) of the different variables at each time to classify evidence on the importance of the different COVID-19 incidence drivers, such that predictors with PIPs above 0.5 are considered as relevant determinants to forecast incidence at that period and specific horizon, and variables below that threshold, as irrelevant ones.^15^

Figure (3) presents the information regarding the PIPs for our different DMA configurations. However, to keep the figure readable, we only present PIPs for the “top predictors”, which are defined as the variables that appear to be part of the forecasting model with a higher frequency over the full time sample considered, and for the three forgetting factors configurations analyzed. Specifically, Figures (3) to (6) plot the PIPs of a variable *k* at a forecasting horizon *h* if its average inclusion probability is in the top quartile of the distribution of the average PIPs for all DMA configurations. To aid interpretation, further note that if the lines in these figures were to be precisely one for any factor, DMA would be using all the models containing this determinant whereas if the lines in these figures were precisely zero, the DMA would completely exclude all the models containing that factor. The interested reader can examine each of the panels in Figures (3) to (6) for any particular variable of interest and horizon. Here we just discuss the main points.

**Figure 3:**
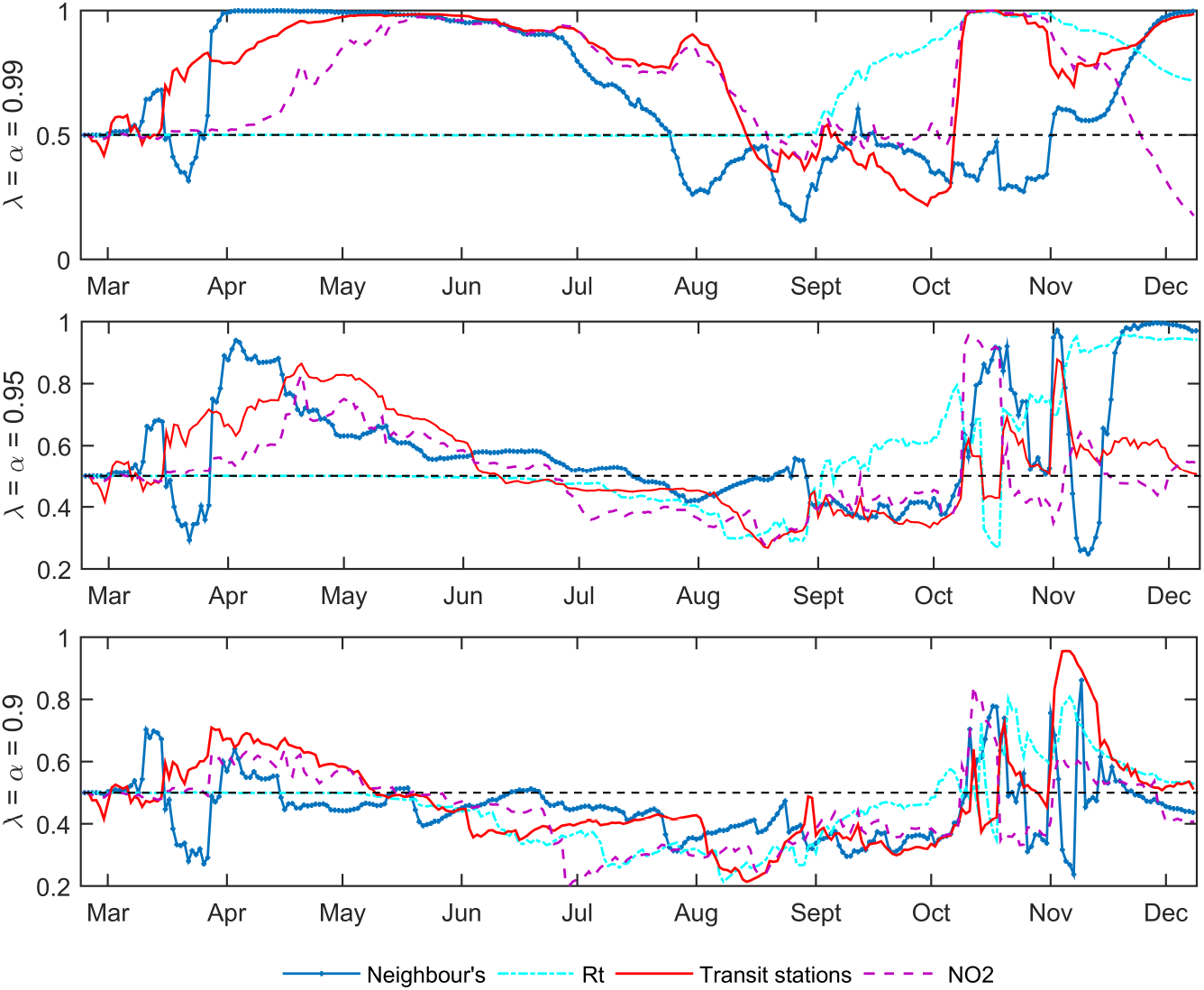
Posterior Inclusion Probabilities for Top Determinants (*h* = 1)

**Figure 4:**
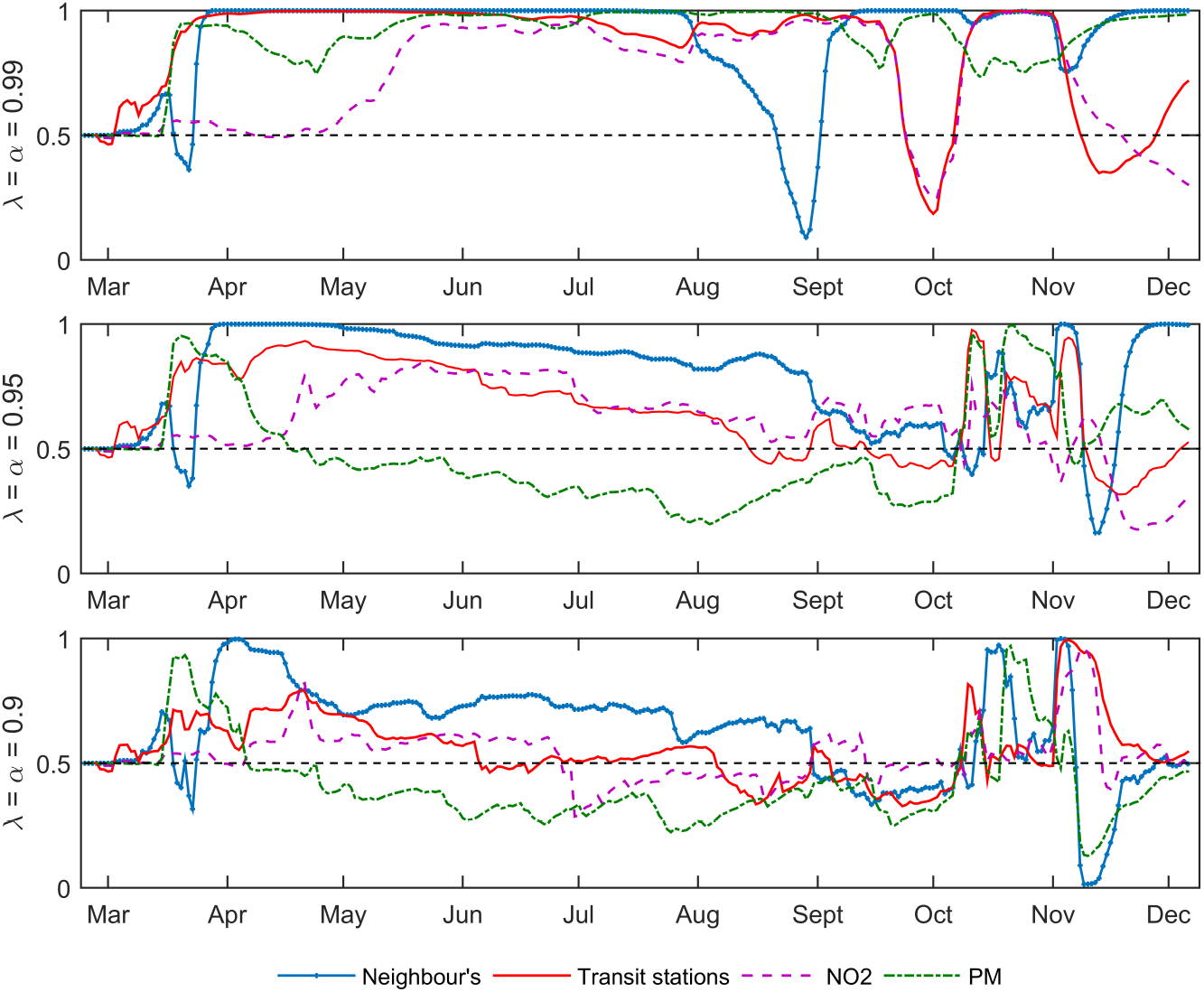
Posterior Inclusion Probabilities for Top Determinants (*h* = 3)

**Figure 5:**
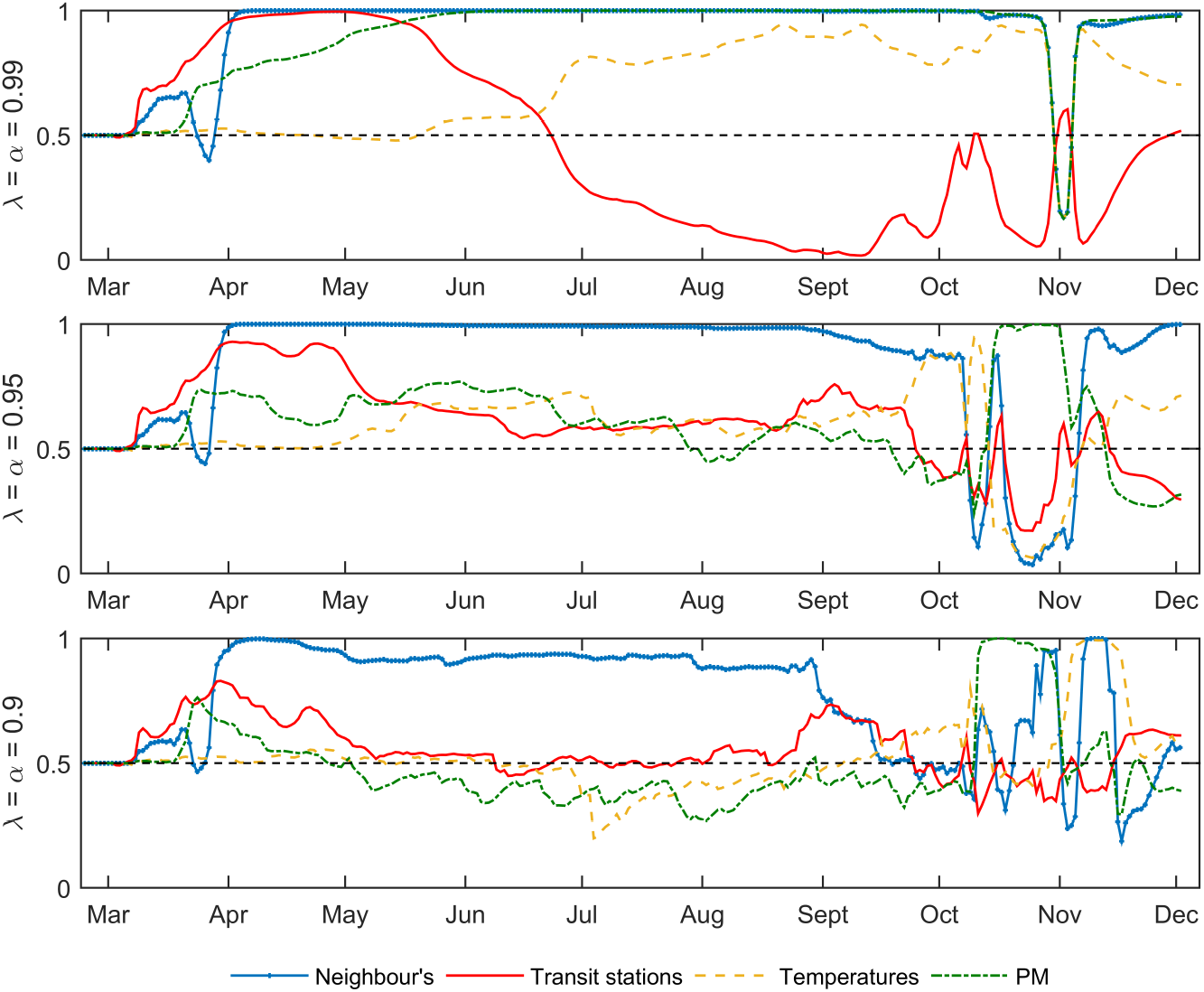
Posterior Inclusion Probabilities for Top Determinants (*h* = 7)

**Figure 6:**
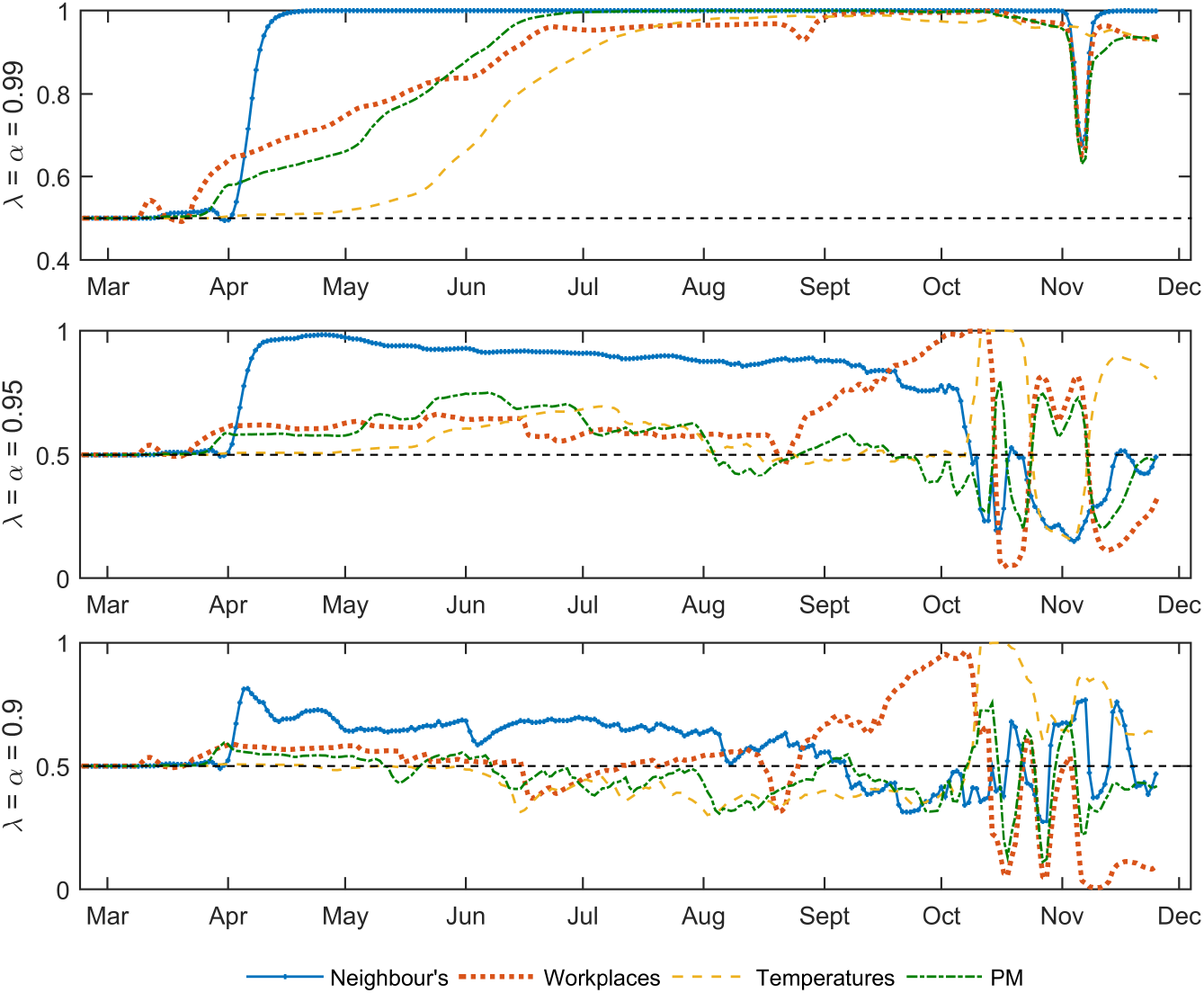
Posterior Inclusion Probabilities for Top Determinants (*h* = 14)

First, for all forecast horizons and DMA configurations we find a strong evidence of model change. That is to say, the set of predictors in the forecasting model is changing over time. Moreover, it can be seen how DMA allows for both gradual and abrupt changes in the role of top predictors. There are many times where the probability associated with a predictor increases or decreases gradually over time (i.e, the gradual increase in the PIPs of temperatures and work mobility when forecasting at *h* = 14 from April to October) but there are also a considerable number of cases where a variable experiments an abrupt change in few periods going from near one to zero (see the PIPs of neighbor’s cases in November 2020 after the implementation of restrictions). This tendency to switch over models is specially remarkable when looking at the changes observed in the PIPs of the top determinants during the second wave.

Second, there is no single predictor that performs well over the entire sample across forgetting factor specifications and horizons. Thus, we find evidence favoring the idea that epidemic predictors are short-lived and relevant for the spread of the disease only in short periods or “pockets of predictability”. Moreover, we do not find evidence of the PIPs of any predictor to display a horizontal trajectory at the 0% for a long period of time, which indicates that DMA is averaging over many models and using many different models with no single variable being consistently dominant, and no single variable being consistently dropped from the forecasting exercise. Therefore, the picture we are finding is one where DMA is averaging over many parsimonious models rather than selecting just a few parsimonious models, and that this set of models is changing rapidly over time.

Third, when analyzing the entire course of the epidemic, the lagged values of the number of new cases in neighboring regions appears to be the most relevant factor, which suggests the relevance of imported cases when driving the spread of the epidemic. In a second level of importance, we find variables included in our set of top drivers. These factors perform well in some periods and horizons, but not in others. This is the case of the mobility to workplaces and stations, the reproductive number, the temperatures or pollution.

As observed in Figures (3) to (6) the human mobility through transit stations is among the top predictors for horizons *h* = 1, 3 and 7 days, with PIPS above the 90% in many configurations and periods. It contributed to forecast new cases until May-June 2020, its importance declined during summer and from September onwards, its predictive importance increased again. On the other hand, when forecasting at the longer horizon of *h* = 14, we find that mobility to workplaces is more relevant than that of transit stations, specially during the period that goes from mid August to mid October, which coincides with the period of children returning to school and adults returning to work. As regards pollution, we find that NO2 pollution is relevant when forecasting *h* = 1 days ahead, whereas suspended particle matter provides valuable insights when forecasting at longer time horizons. Regarding the reproductive number, *R*_*t*_, it behaves as a top predictor but only when performing forecasts one day ahead. As observed, the reproductive number experienced a considerable increase in importance during the second wave, from September onwards. As refers to the climate factors, we observe temperatures do not seem to be relevant when forecasting at short-term horizons, but since the beginning of June, their PIPs experienced a steady increase for *h* = 7 and *h* = 14.

Fourth, none of the policy factors appears to be an overall “top determinant”. The reason is that during the summer period of 2020, which covers more than one third of our sample observations and where incidences where relatively low, these factors were not relevant to forecast incidences at any horizon and experienced PIPS below the 50% threshold. However, as shown in Figures (7) to (10), in some specific periods of key importance from a health policy point of view such, as the beginning of the first wave and the end of the second wave, the predictive performance of these factors was high. During the first months of the epidemic up to May 2020, when forecasting at *h* = 7 and *h* = 14 with the various DMA configurations we find that both, the use of masks and the stringency of the containment policy, registered PIPS above the 50%. This is also the case when forecasting incidences during the period that goes from October to December.

**Figure 7:**
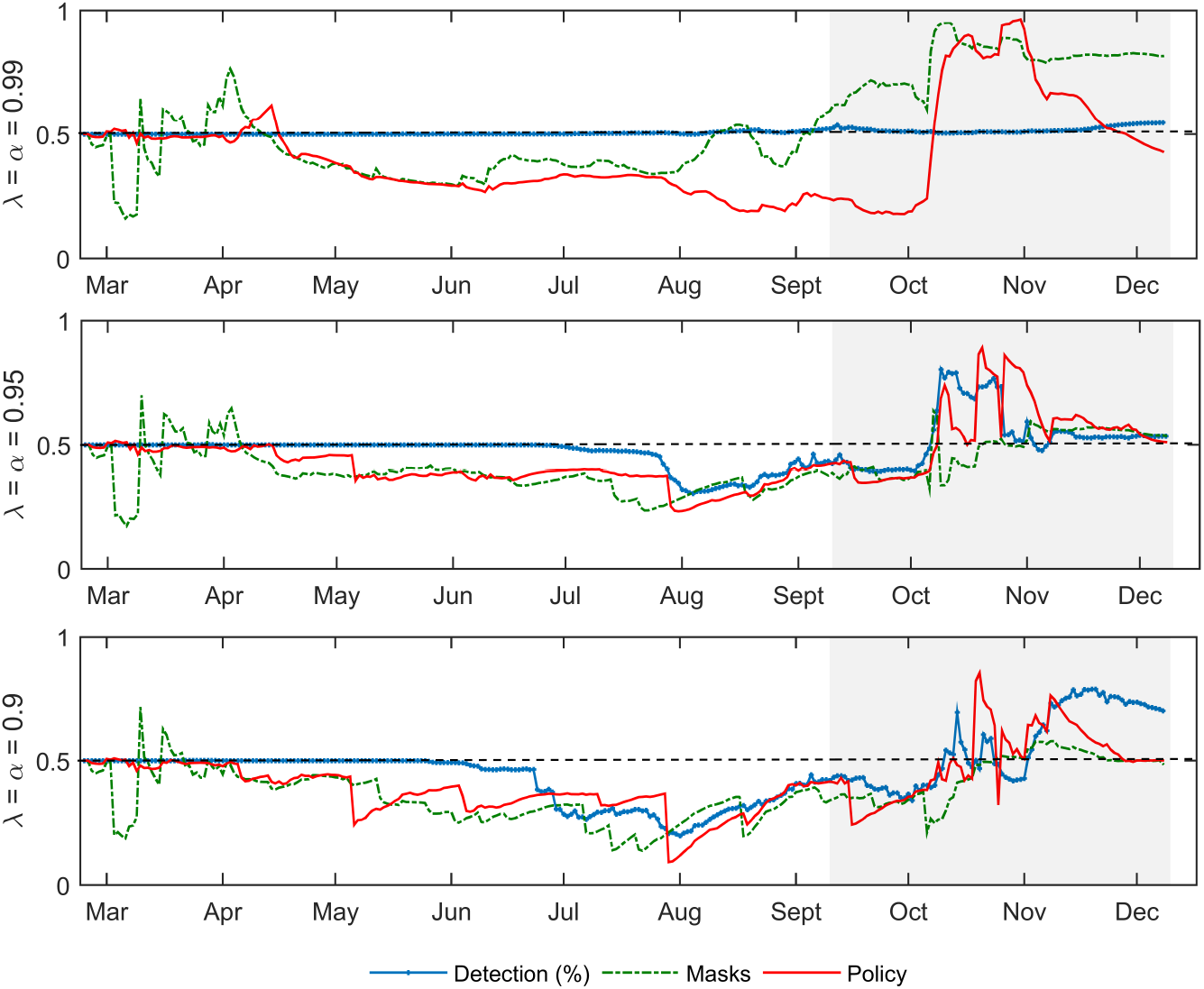
Posterior Inclusion Probabilities for Policy factors (*h* = 1)

**Figure 8:**
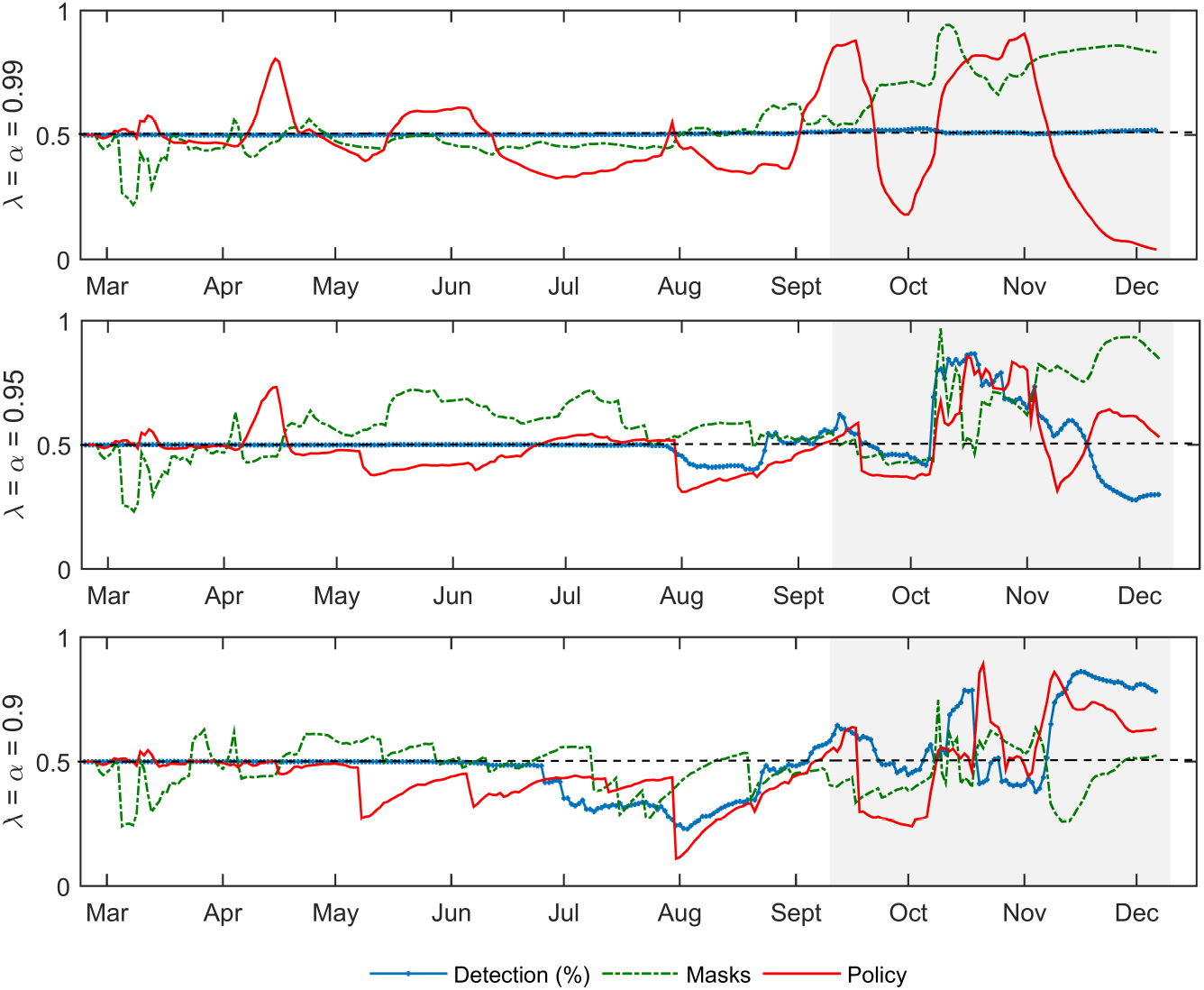
Posterior Inclusion Probabilities for Policy factors (*h* = 3)

**Figure 9:**
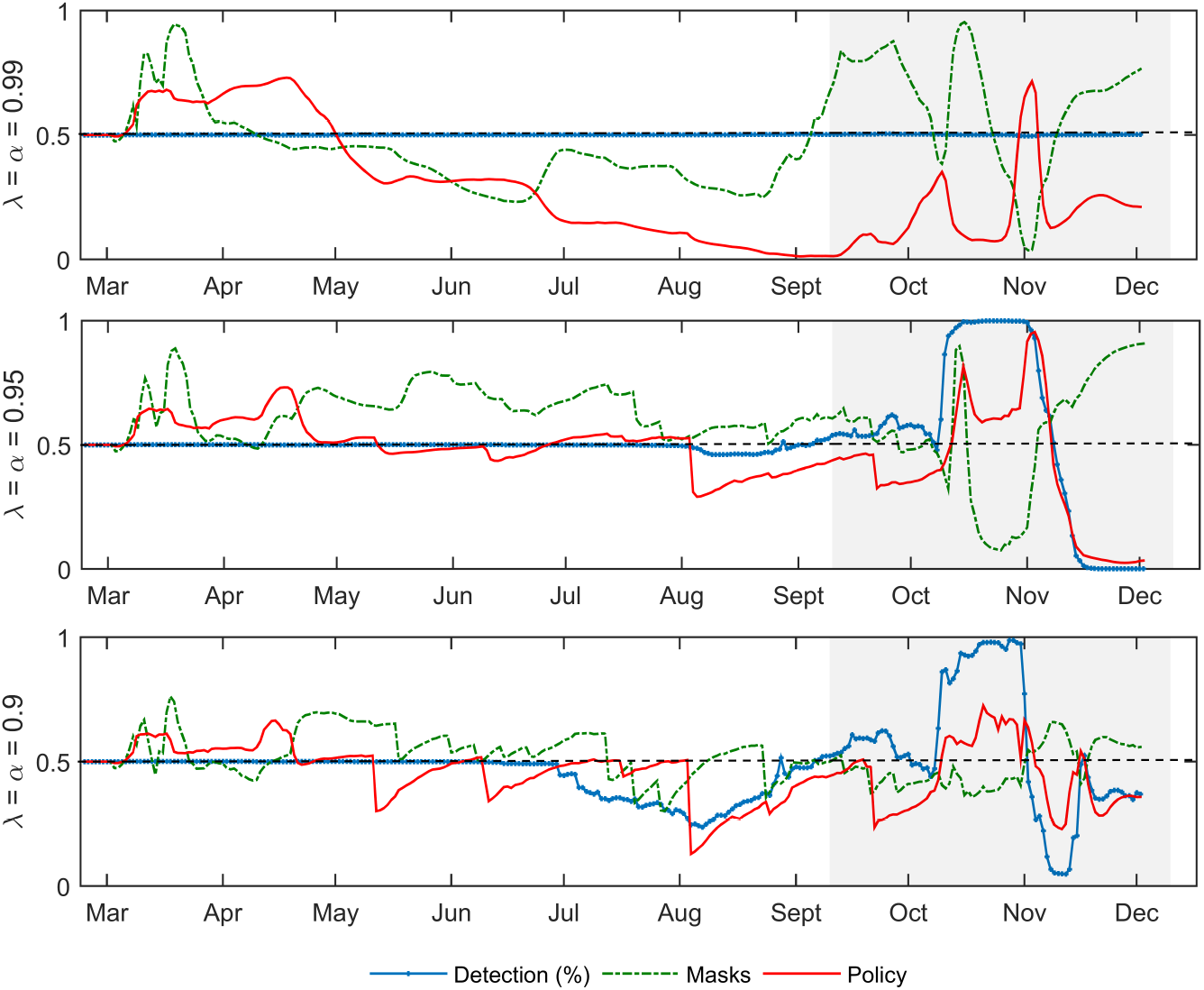
Posterior Inclusion Probabilities for Policy factors (*h* = 7)

**Figure 10:**
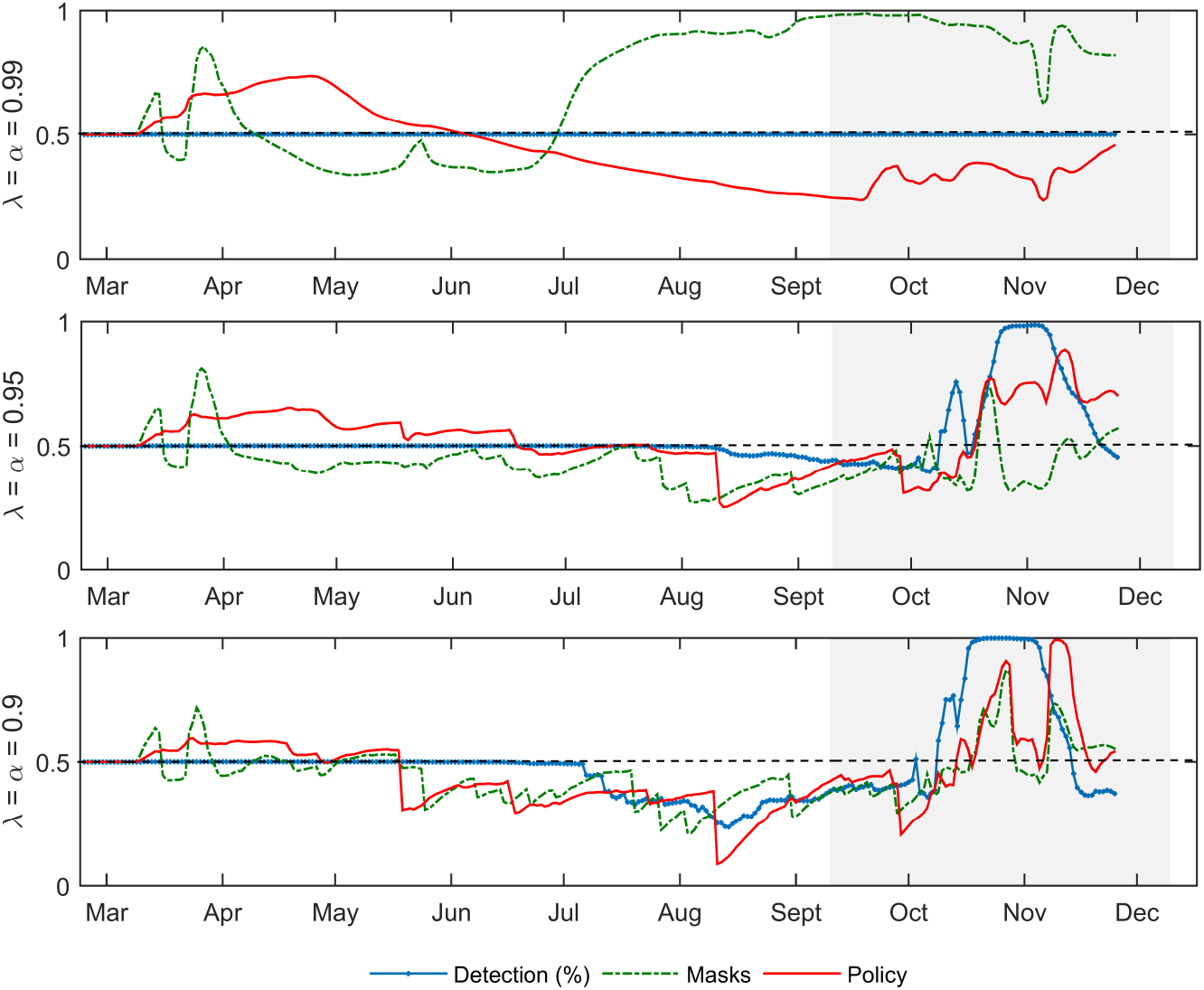
Posterior Inclusion Probabilities for Policy factors (*h* = 14)

However, during the second wave, the specific DMA configuration matters a lot in shaping the variable importance profiles. When using *α* = *λ* = 0.99 the use of masks is the dominant predictor in this category, whereas the share of detected cases receives a steady PIP value of the 50% and the importance of the policy stringency fluctuates. On the other hand, in the DMA configurations of *α* = *λ* = 0.95 and *α* = *λ* = 0.90 we find that from mid October to mid November, the share of detected cases seems more relevant for forecasting than the use of masks, irrespective of the horizon. Under these flexible DMA configurations, the stringency of the policy measures adopted to curve the spread of the COVID during October and November are reflected in higher PIPs, specially when forecasting at *h* = 1 and *h* = 3. Taken together, these results suggest that information on the protective behavior of individuals and epidemic policy measures can be used to forecast incidence, specially before critical turning points.

## 5 Conclusions

This article has investigated the use of DMA and DMS methods for forecasting the time-series of the daily new cases of the COVID-19 pandemic in the Italian region of Lombardy. To our knowledge DMA and DMS have not been used by epidemic modelers and forecasters. This approach developed by Raftery *et al*. (2010) allow for the coefficients in a model to evolve over time, but also allow for the set of predictors used for forecasting to change over time. The alternatives of working with one general model including all potential predictors, or choosing one single parsimonious model are unattractive, given that a good parsimonious forecasting model at some periods could be a bad model at others.

In our empirical analysis, we present evidence indicating the benefits of DMA and DMS. By allowing for both, model and parameter change, DMA and DMS lead to substantial improvements in forecast performance with respect these options. In fact, we find that the DMA forecasting performance is higher the more flexible and quickly adaptable specifications of the forgetting factors, allowing to rapidly capture changes in the transmission speed of the epidemic disease. We also find that the best predictors for forecasting the COVID-19 epidemic are changing considerably over time, and that different factors are automatically picked up by the DMA/DMS depending on the forecasting horizon and the forgetting factor configuration.

When employing different DMA/DMS configurations, we obtain different variable importance trajectories. However, among the set of factors considered, we find that the epidemic dynamics in neighboring regions stand out as the most consistent predictor. In a second level of importance, we find that human mobility intensity in transit stations and workplaces, together with pollution matter pollution and temperatures are of major importance to anticipate the evolution of the epidemic. The general pattern, however, is one where the best forecasting model is changing over time. Indeed, we find that epidemic policy variables such as the stringency of restrictions and bans, the use of masks or the share of detected cases, which do not appear to be top determinants shaping the spread of the disease, due to their relatively low overall inclusion pattern in the forecasting models. However, during the second wave taking place in the fall of 2020, these predictors achieved high posterior inclusion probabilities.

When compared to alternative time-series, machine learning and epidemic modeling frameworks, we find that DMA outperforms all of them in accuracy in terms of density forecasts and interval forecasts. DMA with *α* = *λ* = 0.9 outperforms DMS for *h* = 1, 3 and 7 whereas DMS (*α* = *λ* = 0.99) does the same for *h* = 14. As regards point forecast accuracy, measured by the MAPFE, we find that in the short run the TVP-AR(1)X-SV is the best option whereas the classical SIR model does a good job when in longer horizons. However, the difference between these options and the best DMA/DMS configuration for each horizon is low. Taken together, our results suggest that DMA/DMS methods can greatly contribute to the monitoring and forecasting of the COVID-19 pandemic.

There are some interesting extensions to this research that could be explored to produce more accurate forecasts within the context of DMA/DMS, which in turn could help the decision making of public health officials. One is a sensitivity analysis over a larger set of values for *λ* and *α*. Here we did not explore the optimal configuration of the forgetting factors when minimizing loss functions such as the MAPFE, the ALPL or the MIS. However, forecastability gains in these metrics could be achieved in a more indepth grid-search. A second alternative is to add stochastic volatility in the measurement variance and employ a slower but more exact MCMC estimation procedure. This seems a promising avenue to increase accuracy in forecasts, at least up to one week ahead. Finally, the performance of iterated forecasts relative to that of direct forecasts could be explored. This is because of by design, in our direct forecast setting, parameter estimates used to forecast *h* steps ahead cannot include the most recent information on the linkages between *X*_*t*_ and *y*_*t*_. Given the strong evidence presented here in favor of abrupt parameter and model changes, iterated forecasts using last available information on the relationships between *X*_*t*_ and *y*_*t*_ could potentially reduce forecast errors.

## Data Availability

All the data sources and procedures employed in the construction of the database used in the empirical analysis are detailed in the Appendix A.

https://www.google.com/covid19/mobility/

https://power.larc.nasa.gov/docs/

https://www.eea.europa.eu/themes/air/air-quality-and-covid19

https://github.com/pcm-dpc/COVID-19/tree/master/dati-regioni

## 6 Appendix A: Data

This section details the construction of the database employed in our empirical exercise.

### 6.1 Epidemic Dynamics Variables

- **[1] Reproduction number**

The reproductive number is measure of the instantaneous transmissibility and it is used as a near real-time indicator of epidemic growth. If the *R*_*t*_ *>* 1, the number of cases will increase, if *R*_*t*_ = 1, the disease course will stabilize whereas if *R*_*t*_ *<* 1, there will be a decline in the number of cases. We follow Cori *et al.* (2013) to estimate the *R*_*t*_ as follows:^16^

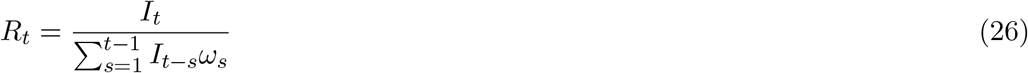

where the numerator is given by the number of new infections generated at time step *t, I*_*t*_, and the denominator is given by the product of the the total infectiousness of infected individuals up to time *t* − 1, weighted by the infectivity function *ω*_*s*_, which is approximated by the distribution of the serial interval. In this study, *R*_*t*_ is calculated specifying an uncertain serial interval distribution (*ω*_*s*_) with a mean of 5.2 (3.7-6.5) days and a standard deviation of 2.9 (1.9-4.9) (i.e, we characterize this distribution using the results of Nishiura *et al*. (2020) and Rai *et al*. (2020)). We employ the statistical package *EpiEstim* developed in R-software to obtain our estimate of the *R*_*t*_ using the default setting of a smoothing sliding window of 7 days.

- **[2] Neighbour’s cases**

Neighbour’s cases are defined as the 7-days moving average of the average 4-nearest neighbour’s incidence. Nearest neighbor’s are calculated using the distance between the centroid of each region vis a vis with Lombardy. Thus, neighbour’s new cases are calculated as:

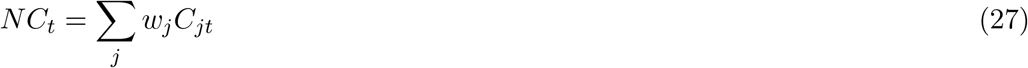

where *w*_*j*_ = 0.25 and *C*_*jt*_ is the daily incidence in region *j* at time *t*. We have experimented with several specifications to define this spatial autoregressive term, finding a 4 nearest neighbor’s performs well in the present context. The four neighboring regions considered are Liguria, Emilia-Romaga, Trento and Bolzano.

### 6.2 Mobility variables

Measurements on mobility come from the Google Mobility Reports (see https://www.google.com/covid19/mobility/). Google mobility reports identify six distinct areas classified by the Google Maps tool: Retail & Recreation, Grocery& Pharmacy, Parks, Workplaces, Transit Stations and Residential areas. These data capture the variation in terms of volume of visitors in the classified places compared to the value of a baseline period. The baseline value represents the median value from the 5-week period Jan 3 Feb 6, 2020 (see Google LLC (2020)).

In our analysis we use a 7-day moving average scaled values to filter out weekend effects (i.e, the baseline period is set to 100 rather than to 0) of the regional mobility data of Lombardy on *(3) Workplaces, (4) Transit stations, (5) Residential areas* and *Parks*.^17^ Each of these categories consist on data on:

- **[3] Work**: mobility trends for places of work.
- **[4] Transit Stations** : mobility trends for places like public transport-hubs such as subway, bus, and train stations.
- **[5] Parks**: mobility trends for places like national parks, public beaches marinas, dog parks, plazas,and public gardens
- **[6] Residential**: mobility trends for places of residence.

### 6.3 Climate variables

Meteorological data is taken from the NASA-Prediction Of Worldwide Energy Resources (NASA-POWER) v8 GIS database (see https://power.larc.nasa.gov/docs/).

All the daily measurement of these climate variables are measured at the coordinates of the regional centroid. The meteorological data-parameters in *POWER Release 8* are based upon a single assimilation model from Goddard? Global Modeling and Assimilation Office (GMAO), whereas the solar based data-parameters in *POWER Release 8* are based upon satellite observations with subsequent inversion to surface solar insolation by NASA? Global Energy and Water Exchange Project-Surface Radiation Budget (SRB) and NASA’s Fast Longwave And SHortwave Radiative project (FLASHFlux) (see https://power.larc.nasa.gov/#resources for details).

Specifically the set of predictors in this group and the formulae used to calculate them are:

- **[7] Mean temperature** 2 meters above the surface in celsius degrees.
- **[8] Mean relative humidity** 2 meters above the surface. The relative humidity (RH) is the ratio of actual partial press of water vapor to the partial pressure at saturation, expressed in percent. RH is calculated as:

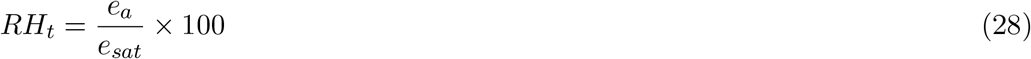

where *e*_*a*_ is the water vapor pressure and *e*_*sat*_ is the saturation water vapor pressure at the ambient temperature *T*_*a*_.

- **[9] Solar radiation**: The daily average amount of the total solar radiation incident on a horizontal surface at the surface of the earth.

### 6.4 Air pollution

Air quality data is taken from the European Environment Agency EEA (see https://www.eea.europa.eu/themes/air/air-quality-and-covid19), which provides daily measurements of NO2, PM25 and PM10 pollutant concentration in (ug/m3) recorded by monitoring stations scattered across cities in European countries. In the region of Lombardy, NO2, PM10 and PM25 measurements are available at various stations different cities, which allows us to measure pollution for each pollutant and city at different spatial locations.

We compute a population weighted average of the daily pollution concentration records for each station located in each city within the region of Lombardy. Since PM2.5 and PM10 indicators display a strong correlation, we use the average of the two indicators of suspended particle matter to produce an overall PM pollution index after applying a max-min normalization to the raw data *PM* 2.5_*raw,t*_ and *PM* 10_*raw,t*_.^18^

- **[10] The PM pollution index** is given by:^19^

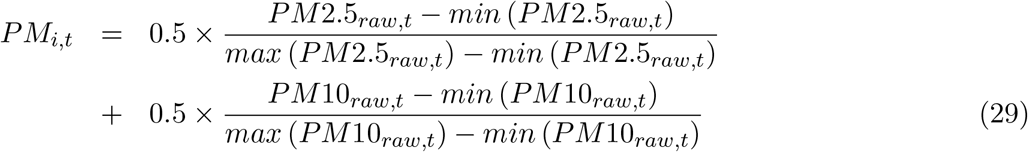

where the *PM* 2.5_*raw,t*_ =Σ _*j*_ *w*_*j*_*PM* 2.5_*j,t*_ and *PM* 10_*raw,t*_ =Σ_*l*_ *w*_*l*_*PM* 10_*lt*_ where *w*_*l*_ is the relative share of the population in city *l* with respect the total population in our sample of cities (i.e, 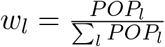. To estimate regional PM2.5 pollution we use available records from the cities of Bergamo, Brescia, Como, Cremona, Lecco, Milano, Pavia and Varese.

For the NO2 and PM10 pollutants the set of cities with measurement stations are: Bergamo, Brescia, Busto Arsizio, Como, Cremona, Lecco, Milano, Pavia, Varese and Vigevano.

Our second indicator to capture the evolution of pollution is a:

- **[12] NO2 pollution index**: measured in (ug/m3). NO2 is a pollutant mainly emitted by road transport. Its aggregation from the city level to the regional level follows that of the PM10 as we the sample of cities with available measurements is the same.

### 6.5 Health Policy and Epidemic Monitoring

- **[12] Health policy containment index**

This is a composite measure based on eleven policy response indicators school closures; workplace closures; cancellation of public events; restrictions on public gatherings; closures of public transport; stay-at-home requirements; public information campaigns; restrictions on internal movements; international travel bans and controls, testing policy and contact tracing, rescaled to a value from 0 to 100 (100 = strictest). The database scores the stringency of each measure ordinally, for example depending on whether the measure is a recommendation or a requirement and whether it is targeted or nation-wide.

- **[13] Detection of cases (%)**

To model the epidemic monitoring by health authorities, we opt for this metric rather than the raw number of tests (or tests per capita) given that a problematic issue with the use of the number of tests as a proxy of the quality of epidemic monitoring is that it is uninformative on the share of cases that are not detected and that could amplify the epidemic later on. We estimate the share of reported of cases following Nishiura *et al*. (2009) and Russell *et al*. (2020) who show that combining a “best estimate” of the lethality and a delay-adjusted case fatality distribution of cases with known outcomes it is possible to obtain daily estimates of the under-reporting of cases in the official statistics. Specifically, we calculate the share of detected cases as:

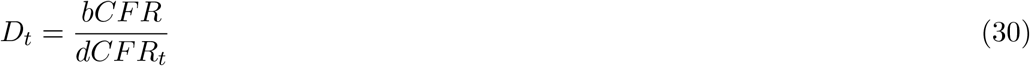

where (i) bCFR denotes the best available estimates of lethality taken from large randomized seroprevalence studies in China, Spain and South Korea, which are in the 1% - 1.5% range (we assume a bCFR = 1.25 %) and 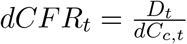 is the delay-adjusted case fatality ratio in *t*. The delay-adjusted case fatality is given by the ratio of the number of daily deaths to *dC*_*c,t*_, which is a correction of the cases accounting for the proportion of cases with known outcomes which is given by:

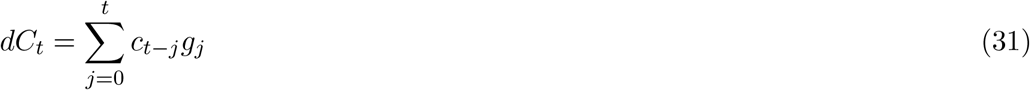

where *g*_*j*_ represents the probability density function between confirmation to death (i.e, we use a lognormal distribution with a mean delay of 13 days and standard deviation of 12.7 days).

- **[14] Use of Masks**

To proxy the use of masks we employ Google Trends search data. Google Trends indexes allow us to capture the relative quantities of web searches through the Google search engine for face masks related keywords, as well as the specified time period, being the values normalized and ranging from 0 to 100. ^20^.

Given the absence of a general pre-defined search category to proxy the use of masks, we select fourteen individual keywords in Italian language that we believe can help to capture variations in the use of face-masks during all the year 2020.

By using the keyword of mask, in Italian language (i.e, “mascherine”) as a benchmark category to obtain trend indexes, we aggregate seven different searches in singular: “mascherina”, “mascherina ffp2”, “mascherina chirurgica”, “mascherina ffp3”, “mascherina KN95” “mascherina con filtro” and “mascherina covid” and seven in plural “mascherine”, “mascherine ffpp2”, “mascherine ffp3”, “mascherine chirurgiche”, “mascherine KN95”, “mascherine con filtro”, “mascherine covid”. We use the sum of all the 14 keyword scores. That is

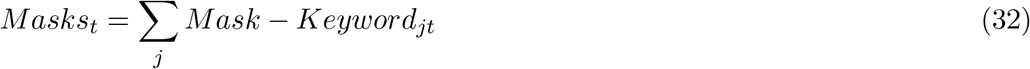

## 7 Appendix B Settings used in competing models

### 7.1 Time Series and Machine Learning models

- TVP-AR1(X)-SV: Time-Varying Parameter Autoregressive Model with Stochastic Volatility. This model is estimated with the efficient MCMC algorithm developed by Chan and Jeliazkov (2009). This is the standard time-varying paramter model regression model used in economics (see Cogley and Sargent (2005)). It consists of the following Equations:

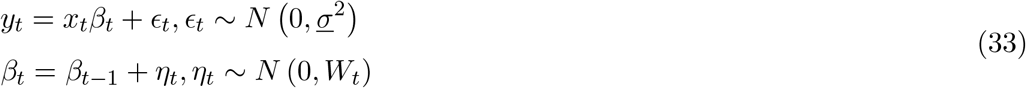

where *x*_*t*_ is a 1 × *K* vector of predictors, _*t*_ and *η*_*t*_ are independent of one another, the measurement variance *<u>σ</u>*^2^ is known, *W*_*t*_ is a diagonal *K* × *K* matrix (i.e, *W*_*t*_ = *diag* (*w*_1*t*_, …, *w*_*K,t*_). The crucial setting that affects the amount of time-variation in the regression coefficients *β*_*t*_ is the prior on state variances *w*_*t*_ which is of the form 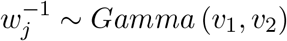. We set *v*_1_ = 3 and *v*_2_ = 20

- BSSVS: Bayesian Stochastic Variable Search Selection. BSSVS is a predictor variable selection method for Bayesian linear regression that searches the space of potential models for models with high posterior probability and averages the models it finds after it completes the search. The static variable selection prior of George and McCulloch (1993) developed for the constant parameter regression using MCMC is of the form:

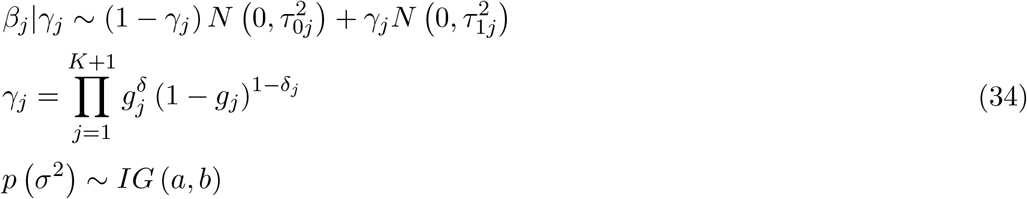

for *j* = 1, …, *K* where *a, b* and *g*_*j*_ are fixed prior hyper-parameters and 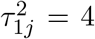 and 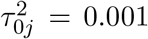 By setting 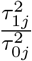 to a large number, the latent binary variables *γ*_*j*_ govern which one of the normal distributions above is active. When *γ*_*j*_ = 0 because of 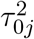 is very small we shrunk the variable *j* corresponding parameter towards 0 whereas if *γ*_*j*_ = 1, the prior exerts little influence on the posterior. We set *a* = *b* = 0.01 and *g*_*j*_ = 0.5. Therefore the prior probability of inclusion of each variable *X*_*j*_ is the 50%.

- BAG: Bagging. Bagging stands for “Bootstrap aggregating”. With the bagging algorithm we first re-sample our data *B* times, with replacement blocks of size *m*. For each pseudo-generated data set we estimate with ordinary least squares using the Newey and West estimator of the covariance with lag truncation parameter int *T* ^1*/*4^. In each draw we select the optimal model using only those predictors that have t-statistics larger than a threshold *c*^*^ in absolute value. We forecast with the optimal model, and the bagging forecast is obtained as the average of all forecasts over the *B* Bootstrap replications. We set *B* = 1000, *m* = 1 and *c*^*^ = 1.965
- PLS: Partial Least Squares (PLS) is a method that originated in chemometrics (see De Jong (1993)). It allows to estimate factors that are extracted with reference to the variable to be predicted (target variable). A key difference with principal components is that the later only maximize the variance explained by the large dataset, and may not be optimal for prediction of the target variable. While more elegant methods have been proposed recently for prediction, the PLS is undeniably a good benchmark for assessing whether we can improve on the information content of simple principal component estimates. We use again the MATLAB function “plsregress” available in the Statistics and Machine Learning Toolbox, and we extract fifteen factors from our dataset.

### 7.2 Epidemic Models

- Phenomenological models

We consider three distinct phenomenological models of epidemic growth. The first model is the (i) Generalized Logistic Growth Model (GLGM) which is given by the following ordinary differential equation

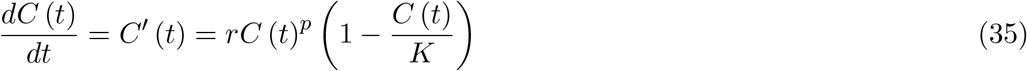

where *C* (*t*) is the cumulative cases at time *t, r* is the early growth rate and *K* is the carrying capacity. This specification extends the simple logistic growth model with a scaling of growth parameter *p* ∈ [0, 1] that accommodates sub-exponential growth patterns. Our second candidate is the (ii) Generalized Gompertz Growth Model (GGoM) which is given by:

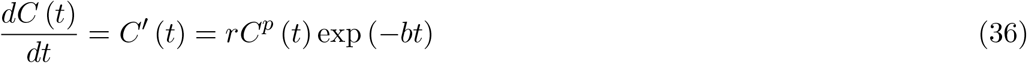

where *b >* 0 describes the exponential decay of the growth rate *r*. Finally, (iii) the Generalized Richards Growth Model (GRGM) is given by:

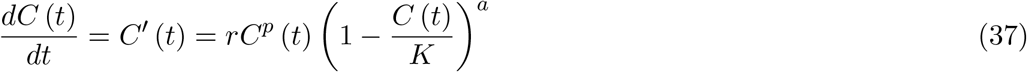

We approximate the solution of the ODEs described above using the Runge-Kutta (4,5) iterative numerical method given the initial condition, *C*_0_ using the ode45 solver of Matlab. Once we have the numerical solution of the ODE, we estimate the best-fit model solution to the reported data using weighted nonlinear least squares fitting (WNLSQF).

That is we fit the evolution of contagions by minimizing: ^21^

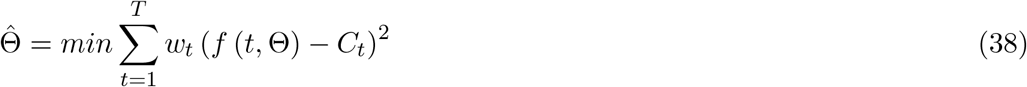

where *w*_*t*_ = *w*_*t*−1_ (1 − *α*) and *w*_0_ = *α* (i.e, with a higher *α* value we attribute more weight to the most recent data). This simple exponential smoothing regulates the rate at which the weights decrease by setting *α* ∈ [0, 1]. In our empirical analysis we set *α* = 0.5. Parameter uncertainty is investigated by means of bootstrap methods by sampling from a Poisson distribution. Using the best model fit 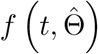 we generate S-times replicated simulated datasets denoted by 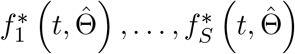 by drawing from:

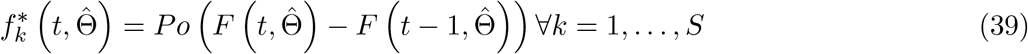

where 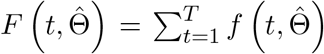. We then re-estimate the parameters for each of the S-simulated realizations given by 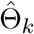 These re-estimated parameters are used to characterize the empirical distribution of 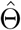 (see Chowell (2017) pp 385-386 for details). Finally, forecasts are generated by propagating the estimated model uncertainty given by 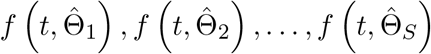 *S* in time by a horizon of *h* time units as follows:

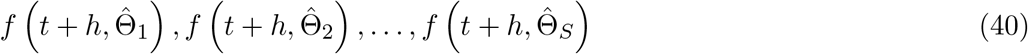

Therefore, we forecast the entire uncertainty of the system using the uncertainty associated with the parameter estimates which allows us to construct the 95% confidence intervals.

- SIR. Susceptible-Infected-Removed model. The SIR model classifies individuals in the compartment as one of three classes: susceptible (S), infectious (I), and recovered or removed (R). Infectious individuals spread the disease to susceptible individuals at rate *β* and remain in the infectious class for a given period of time known as the infectious period before moving into the recovered (or removed) class at rate *γ*. Individuals in the recovered class are assumed to be immune for an extended period (or removed from the population). For the total population *N* = *S* + *I* + *R*, the dynamical system describing the SIR equations is given as:

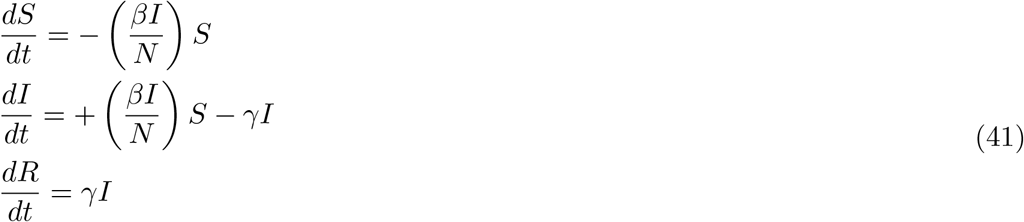

To connect the model with the data, we will use the following measurement equation: 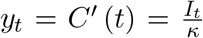, where 1*/κ* is a combination of the reporting rate, the asymptomatic rate, and the total population size.

We fit the SIR model by means of Maximum Likelihood using the “fminsearch” algorithm of unconstrained nonlinear optimization in Matlab assuming a Poisson data generating process for the incidences and providing the following initial parameter guesses *β*_0_ = 0.4 and *γ*_0_ = 0.25, *κ*_0_ is set to 80,000. With our fitted parameters values in hand, in sample fitted trajectory of infections *Ît* and *y*_*t*_ are obtain using the ode45 solver after passing initial conditions I(0), R(0) and S(0). Uncertainty is investigated by means of bootstrap methods by sampling from a Poission distribution as in the context of phenomenological models. Again, each re-estimated parameter draw is propagated forward to produce out-of-sample forecasts and derive confidence bands.

## Footnotes

1 The key advantage of phenomenological models over compartmental ones, relies on the fact that they provide an empirical approach to the analysis of the expected trajectory of the disease, without a specific basis on the physical laws or mechanism that give rise to the observed patterns in the data (Chowell, 2017).

2 Although restricted to the context of phenomenological models, to the best of our knowledge, only Chowell *et al*. (2020) have developed an empirical approach to ensemble epidemic model forecasts within the context of various phenomenological models.

3 Rare exceptions using time-varying parameter compartmental models in the field of epidemics are Cakmakli and Simsek (2020) or Ferrari *et al*. (2020).

4 An example of issues when registering and compiling data in a similarly affected, developed and decentralized countryis that of Spain. As explained by the New York Times (2020), Spain has been implementing a number of methodological changes for logging deaths and cases, leading to fluctuations in its statistics and frequent revisions of data.

5 For details on the estimation of the true epidemic size and the % of detected cases see the Appendix A.

6 As explained by Raftery *et al*. (2010), if *W*_*t*_ = 0 for *t* = 1, …, *T* then *θ*_*t*_ will be constant, so that this model nests fixed parameter linear regressions parameters. It should be noted that ultimately, the variation in the regression coefficients captured by *θ*_*t*_ depends on the data.

7 Note that the employment of Equation 7 instead of 6 is equivalent to set 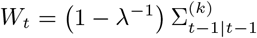.

8 The intuition of this factor is that for the estimation of the parameters in *t*, the observations that are *i* periods old, receive a weight *λ*^*i*^ and the amount of data used for the estimate (or the window size) is *h* = 1*/* (1 − *λ*). When *λ* = 1, *θ*_*t*_ will be constant over time whereas with *λ* →0 only the most recent information is used for forecasting, or equivalently we allow for large structural breaks can occur.

9 The interpretation of *α* becomes clear if we rewrite Equation 11 as follows: 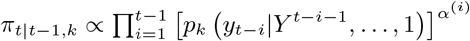 where it can be seen that values *α* →1 will imply that *π*_*t*|*t*−1,*k*_ will be larger at time *t* if it forecasts well in the past.

10 The log predictive density for the *h*-step ahead forecast is the logarithm of the *h*-period extension of this.

11 We use the value of the median forecast trajectory at each *h* as our point forecast, *ŷ*_*τ*|*τ*−*h*_

12 However, this is at the cost of not using all available information when producing our real-time forecasts which decreases the quality of the forecast. To clarify this, note that when forecasting in real time at date *t, h*-steps ahead, we use parameter estimates that are *h*-periods old 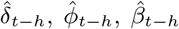 to produce *ŷ*_*t*+*h*_: 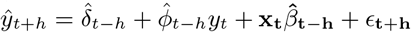 This contrasts with the common practice of previous DMA studies of Bork and Møller (2015); Koop and Korobilis (2012), Naser (2016) or Drachal (2016) among others, where direct forecast performance metrics are derived from an estimate of *ŷ*_*t*+*h*_ obtained using 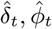 and 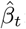 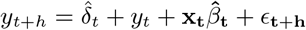 However, when forecasting in real time the estimates of 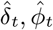 and 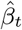 to predict future incidences *ŷ*_*t*+*h*_ can only be used when implementing iterated forecasts and not in the context of direct forecasts. We believe the issue of direct/iterated forecasting for *h >* 1 is an interesting area of research in the context of the COVID-19, as the evidence and arguments on the superior performance of direct vs iterated forecast provided by Marcellino et al. (2006) are based on fixed-parameter AR(1)-X models.

13 Other than the estimation of the TV-AR-SV which relies on the exact likelihood function to implement the Monte Carlo Markov Chain estimation procedure, the stochastic volatility specification is different from the TVP-AR with forgetting factor in that it allows the measurement variance *V*_*t*_ to follow a log stochastic volatility specification and it restricts the state covariance matrix *W*_*t*_ to be constant.

14 However, a problematic issue with them is that they cannot fit properly multiple epidemic waves. For this reason, we estimate them in each wave separately and average their performances by the relative sample size in each wave.

15 From a Bayesian perspective, predictors with PIPs higher than others, reflect a higher importance as they are more likely to be part of the data generating process, and as such, they can be considered as a relevant factor forecasting the evolution of the pandemic. Equivalently, PIPs are the weight used by DMA attached to models which include a particular predictor.

16 We opt for this approach instead of the alternative of Bettencourt and Ribeiro (2008) or **?** because these approaches either require data from after time *t*, or rely on structural assumptions that if are not satisfied, yield biased estimates of the *R*_*t*_ (see Gostic *et al*., 2020 for a critical review on the measurement methods of *R*_*t*_).

17 We do not include the mobility data time series for the categories of Retail & Recreation and Grocery& Pharmacy as they are highly correlated with the Workplace and Transit Stations indicators to avoid collinearity issues.

18 The sources of this pollutant are varied, including the combustion of fuel for the heating of residential, commercial and institutional buildings, industrial activities and road traffic.

19 Using other procedures of data reduction such as principal components gives a weighting scheme of 0.49 and 0.51.

20 The maximum value of each region-keyword specific index is assigned to the peak of the respective time series during that period

21 In Matlab (The Mathworks, Inc.), two numerical optimization methods are available to solve the nonlinear least squares problem: The trust-region reflective algorithm and the Levenberg-Marquardt algorithm. We employ the trust-region-reflective since we impose bound constraints on the parameter values. Moreover, the ode solvers need a guess on the parameters *r*_0_, *p*_0_, *b*_0_, *a*_0_ and *K*_0_ to initialize the search. We set *r*_0_ = *p*_0_ = *a*_0_ = 0.5, *K*_0_ = *C*_*t*_ when implementing the GRGM and the GLGM. The parameter constraints imposed in these contexts are *r* ∈ [020], *p* ∈ [0, 1], *a* ∈ [0, 20] and *K* ∈ [*K*_0_, 20*K*_0_]. For the GGoM we set 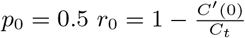 and 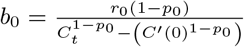

